# The Michael Mason Prize: Development and feasibility testing of a complex intervention to improve adherence to fracture prevention medicine using a person-centred approach

**DOI:** 10.1101/2025.03.21.25324401

**Authors:** Laurna Bullock, Natasha Tyler, Emma M Clark, Simon Thomas, Christopher Gidlow, Ida Bentley, Ashley Hawarden, Nicola Peel, Celia L Gregson, Stuart H Ralston, Joanne Protheroe, Janet Lefroy, Terence W O’Neill, Christian Mallen, Clare Jinks, Zoe Paskins

## Abstract

**Objectives:** The **i**mproving uptake of **Fra**cture **P**revention treatments (iFraP) intervention consists of an osteoporosis decision support tool (DST), clinician skills training, and information resources, to improve shared decision-making (SDM) about, and uptake of, osteoporosis medicines in Fracture Liaison Services (FLS). This paper details the development and feasibility of the prototype intervention.

**Methods:** Intervention development was underpinned by (1) theories of SDM, medicines adherence, and behaviour change; (2) integrated findings from six discrete development studies; and (3) extensive public contributor, patient, and clinician contribution to identify key ‘needs’ for the intervention to address and the intervention’s content, functionality and scope.

Feasibility testing was conducted at one FLS. Intervention consultations were observed and audio recorded, and interviews completed with FLS clinicians and patients to explore perceived acceptability and feasibility.

**Results:** Intervention development identified patient and clinician unmet needs for personalised and evidence-based information about osteoporosis, its consequences, and its treatment within and after FLS consultations, to facilitate clinical and SDM about medicines. The prototype intervention was designed to meet identified needs and overcome barriers to use.

Clinicians delivered the prototype iFraP intervention in 10 consultations with consenting patients. Findings demonstrated that the intervention was acceptable and feasible to deliver, with potential to improve patient outcomes. The intervention was refined to support implementation.

**Conclusion:** The multi-facilitated approach to intervention development and testing ensured that the iFraP intervention was acceptable and feasible for use in UK FLS to support SDM about osteoporosis medicines. The iFraP trial will evaluate implementation, and cost and clinical effectiveness.

**Key messages:**

- Verbal and written communications concerning osteoporosis are currently confusing, technical and impede decision-making and medicine uptake
- Key barriers to implementing shared decision-making and decision aids include clinician goals, professional roles and contextual factors
- A decision tool with clinician training and information resources was co-designed, acceptable, and addressed identified needs and barriers

## Introduction

Osteoporosis is a common condition of weak bones characterised by a reduction in bone strength which increases propensity to fragility fracture. There are over 500,000 fragility fractures each year in the UK, costing the NHS £4.7 billion annually[1] and fragility fractures are the fourth leading cause of disability among non-communicable diseases in Europe[1].

For people with increased fracture risk, evidence-based osteoporosis medicines (anti-resorptive and anabolic treatments) are recommended for the treatment of osteoporosis by the National Institute for Health and Care Excellence (NICE)[2]. Among those who are recommended osteoporosis medicines by specialist Fracture Liaison Services (FLS), only 12% of people are reported as taking them 1-year post-fracture[3].

Shared decision-making (SDM) has the potential to support osteoporosis medicine adherence[4,5], by ensuring the treatment is a good ‘fit’ for the patient[6]. NICE recommend all clinicians support SDM, as a key component of person-centred, personalised care, and use good quality decision support tools (DSTs) where available[7,8]. Across a range of conditions, DSTs have been shown to increase patient knowledge and participation in decision-making, reduce decisional conflict, and improve the accuracy of risk perception[4].

The **i**mproving uptake of **Fra**cture **P**revention treatments (iFraP) intervention was conceptualised in response to a patient priority setting exercise calling for increased accessible information from health professionals about the safety and benefit of osteoporosis medicines[9]. The iFraP intervention principally consists of a DST and enhanced clinical skills training, aiming to improve SDM about, and uptake of, osteoporosis medicines. This paper describes the development and feasibility of the prototype intervention.

## Methods

### Overview and intervention development approach

The Medical Research Council’s (MRC) guidance for developing and evaluating complex interventions was used as an overarching framework[10]. We drew on the three-step implementation of change model[11], which asks three pragmatic questions about the intervention and how it interacts with contextual factors (defined as: “any feature of the circumstances in which an intervention is conceived, developed, implemented and evaluated”[10]) and future implementation: 1) where do we want to be? 2) where are we now? and 3) how do we get there? To answer these questions, we used an evidence and theory-based intervention development approach, working in partnership with public contributors and stakeholders[12]. An overview of the development process is shown in Figure 1, which was dynamic, flexible and iterative. The protocol for the iFraP development work and the findings of each development study are described in detail elsewhere[13–18]. This paper highlights how each study influenced intervention development (reported in accordance with the GUIDED guidelines[19]), and the feasibility findings.

**Figure 1:**
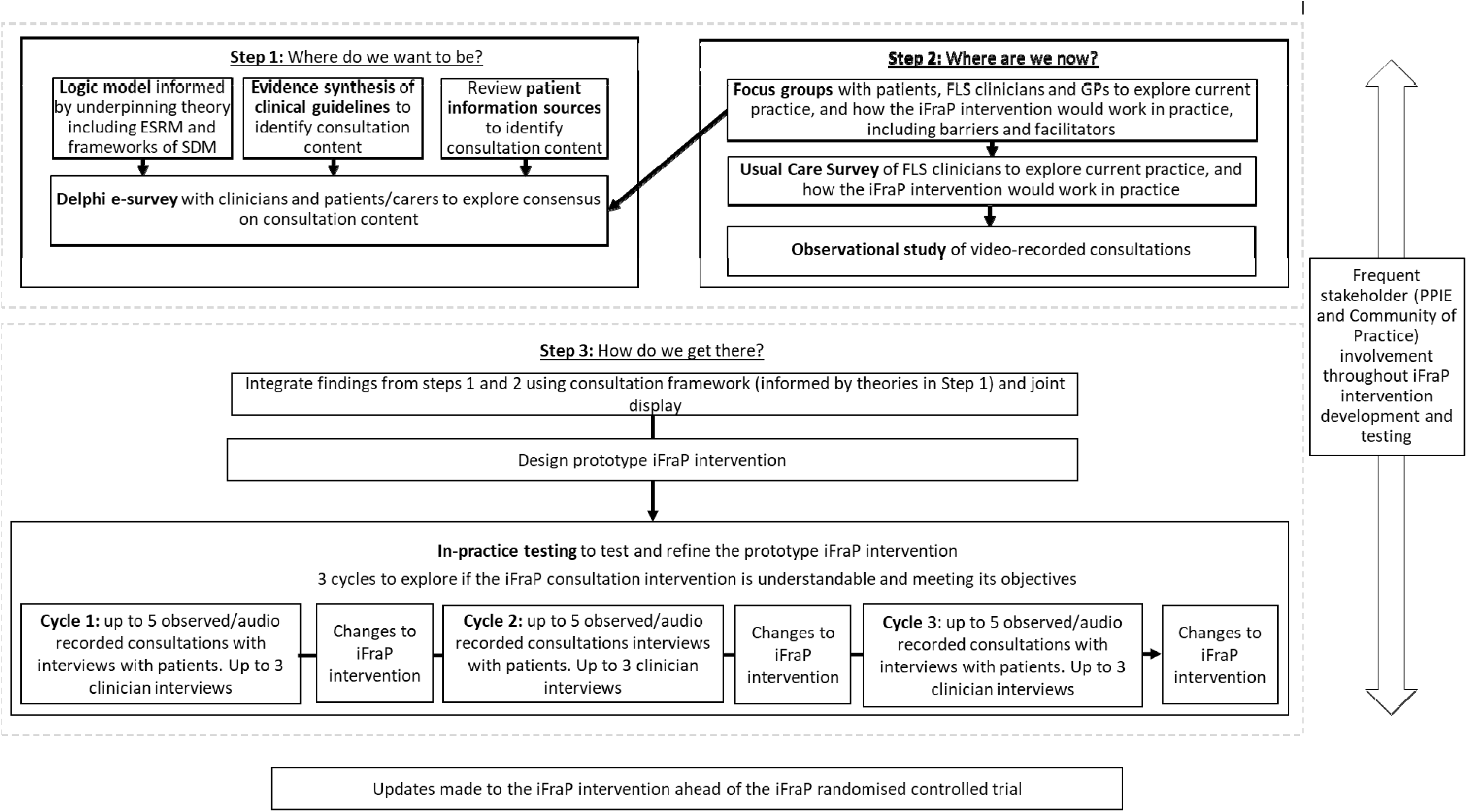
Overview of iFraP development process and methods^1^ ^1^ESRM Extended Self Regulatory Model; SDM Shared Decision Making, FLS Fracture Liaison Service, PPIE Patient and Public Involvement and Engagement; GP General Praactitioner

### Target population and intervention context

The iFraP intervention was developed to be used by UK FLS clinicians and patients. FLS (or sometimes called ‘Fracture Prevention Services’) are usually allied to either rheumatology, endocrinology or geriatric medicine specialties. In this paper, FLS ‘clinicians’ include nurses and allied health professionals who enact secondary fracture prevention by systematically identifying adults aged ≥50 years with fragility fractures, conducting bone health assessments and providing treatment recommendations[20].

### The iFraP team and co-design approach

The iFraP intervention development and feasibility testing research team included expertise from clinical practice, applied health research, and the third sector. The team had specialised knowledge and experience in developing and testing complex interventions, software development, behaviour change, health literacy, medicines adherence and communication skills.

A Community of Practice (CoP) brought together stakeholders with a common concern or interest with the aim of improving and learning to do better through regular group interaction[21]. CoP members included FLS clinicians, General Practitioners (GPs), osteoporosis specialists, commissioning representatives, public contributors (people with lived experience of osteoporosis and/or family members), representatives from the Royal Osteoporosis Society (ROS) and Health Literacy UK. Six CoP meetings supported decision-making and codesign of intervention content and structure.

Public contributors also attended regular Patient and Public Involvement (PPI) meetings as well as study team meetings, steering group meetings, and analysis discussions. An public contribution ‘impact log’ demonstrates enhanced accessibility and inclusivity of the intervention and the research design (see Supplementary Table S1).

### Step 1: “Where do we want to be?” Make a concrete proposal for change, and develop the content and format of the consultation intervention

#### Programme theory and underpinning theoretical framework

The intervention programme theory (or ‘logic model’) details the iFraP intervention resources, hypothesised mechanisms and outcomes, and contextual factors and was refined iteratively throughout intervention development (see Supplementary Figure S1).

The Extended Self-Regulatory Model[22], incorporating Leventhal’s Common-Sense Model (CSM) of Illness[23] and the Perceptions and Practicalities Approach (PaPA)[22], provided a theoretical foundation for the iFraP intervention by explaining how a person’s beliefs about osteoporosis and treatments are linked to their decision and behaviour to adhere to medicine. Leventhal’s CSM of Illness outlines beliefs that help people to ‘make sense’ of their illness. For example, it’s common for people to not understand their bone health (illness coherence) and have low perceptions about how controllable osteoporosis is (illness controllability)[24]. The PaPA (which incorporates the Necessity Concerns Framework[25]) informs the UK’s NICE guidance on medicines adherence[26], postulating that when patients make decisions about taking a prescribed medicine, they weigh up their perceived need for the medicine (necessity beliefs) against concerns about the medicine (concern beliefs), and practicalities of taking the medicine.

To inform our approach to SDM, we used the Ottawa Decision Support Framework[27], a model for designing decision aids, and the SPIKES protocol[28] which is commonly used to teach person-centred communication skills. Core shared features of both frameworks include the focus on eliciting and addressing the patient’s illness and medicine perceptions (described above), building on the patient’s existing knowledge, and establishing SDM preferences.

Few SDM interventions address the needs of adults with lower health literacy[29]. Our approach incorporated evidence-based health literacy techniques to:

- ensure that information shared was understandable (e.g. principles of universal precautions for health literacy, Chunk and Check[30])
- improve accessible risk communication, including the use of simple frequencies, absolute rather than relative risks, and positive and negative framing[31]
- check patient understanding using techniques such as TeachBack[32])

#### iFraP intervention development studies

Three intervention development studies guided the content and format of the iFraP consultation intervention. Each study’s methods are briefly described below, with further detail reported in their respective publications.

#### Evidence synthesis of existing osteoporosis decision aids[15]

A core element of complex intervention research is identifying existing interventions[10]. We conducted a systematic review and environmental scan to identify existing osteoporosis decision aids and assess their quality and efficacy and we discussed results with public contributors.

#### Evaluating existing patient information about osteoporosis[13]

We purposively sampled online UK patient information resources about osteoporosis. We examined their quality, including readability. We extracted frequently used phrases and descriptors about osteoporosis and osteoporosis medicines and worked with the CoP to review this terminology and identify optimum understandable and accurate language to use in the intervention.

#### Delphi e-survey gaining consensus on intervention content[17]

A modified Delphi survey provided structure and content for the iFraP intervention by determining consensus on tasks for UK practicing clinicians in a model FLS consultation. Statements were generated by reviewing UK clinical guidelines for the assessment and management of osteoporosis/fragility fractures; the conduct of the consultation to enhance patient experience; and medicine adherence. Additional statements were incorporated from theories and frameworks of SDM, iFraP development studies, and CoP and public contributor discussions.

### Step 2: “Where are we now?” Understand the current clinical context including barriers and enablers to behaviour change

Four intervention development studies were conducted to understand UK FLS and barriers and enablers to using a DST and implementing SDM in osteoporosis consultations. Each study’s methods are briefly described below.

#### Qualitative study with patients and clinicians[16]

To understand current FLS practice and determine barriers and enablers to using a DST to facilitate SDM conversations in FLS consultations, we conducted focus groups and semi-structured interviews with patients who had consulted in FLS, FLS clinicians, and GPs. Data collection was facilitated by stimulus material in the form of images of existing osteoporosis DSTs and Cates plots (presenting fracture risk in simple frequencies).

Inductive framework analysis[33] was mapped onto the Theoretical Domains Framework (TDF) domains[34] to help understand barriers and enablers to using a DST and improving SDM about osteoporosis medicines. To consider and optimise iFraP intervention acceptability, we also used the Theoretical Framework of Acceptability (TFA) as an overarching framework[35].

#### Video-recorded osteoporosis consultations[36]

We supplemented our development work with an observational study (secondary analysis) of video-recorded consultations to explore how beliefs about osteoporosis and osteoporosis medicines are elicited and addressed by GPs, using a bespoke coding tool.

#### Exploring how COVID impacted the clinical context: E-survey of FLS usual care[14] and secondary analysis of qualitative data[37]

The iFraP DST was originally conceptualised for use in face-to-face appointments. The COVID pandemic accelerated the widespread adoption of remote consultations[38]. In response to this changing clinical context, we conducted two additional studies not initially part of the development protocol:

1. A usual care e-survey of UK FLS practice to quantify the extent of remote consulting and reassess how the iFraP intervention would function and interact with the changing FLS context[14].
2. A secondary analysis of focus group and interview data[16] to explore the acceptability of, and preferences for, remote consulting, using the TFA as a deductive framework[37].

### Step 3: “How do we get there” - Develop a strategy to change behaviours, by designing and refining the prototype into a draft intervention and feasibility testing

#### Integration of findings

Findings of the development studies were summarised using joint displays which were interrogated to identify meta-inferences (see Table 1).

**Table 1.**
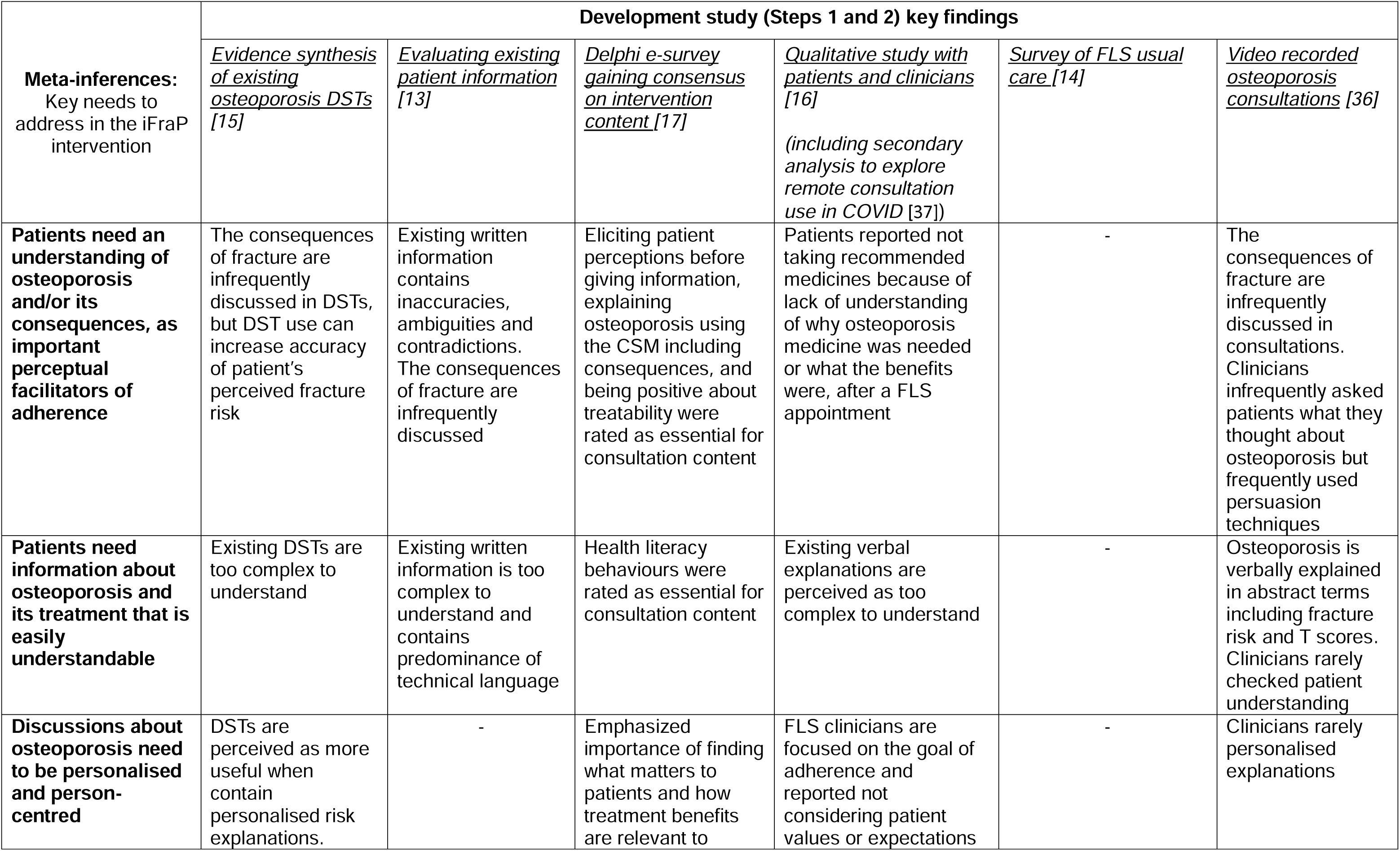
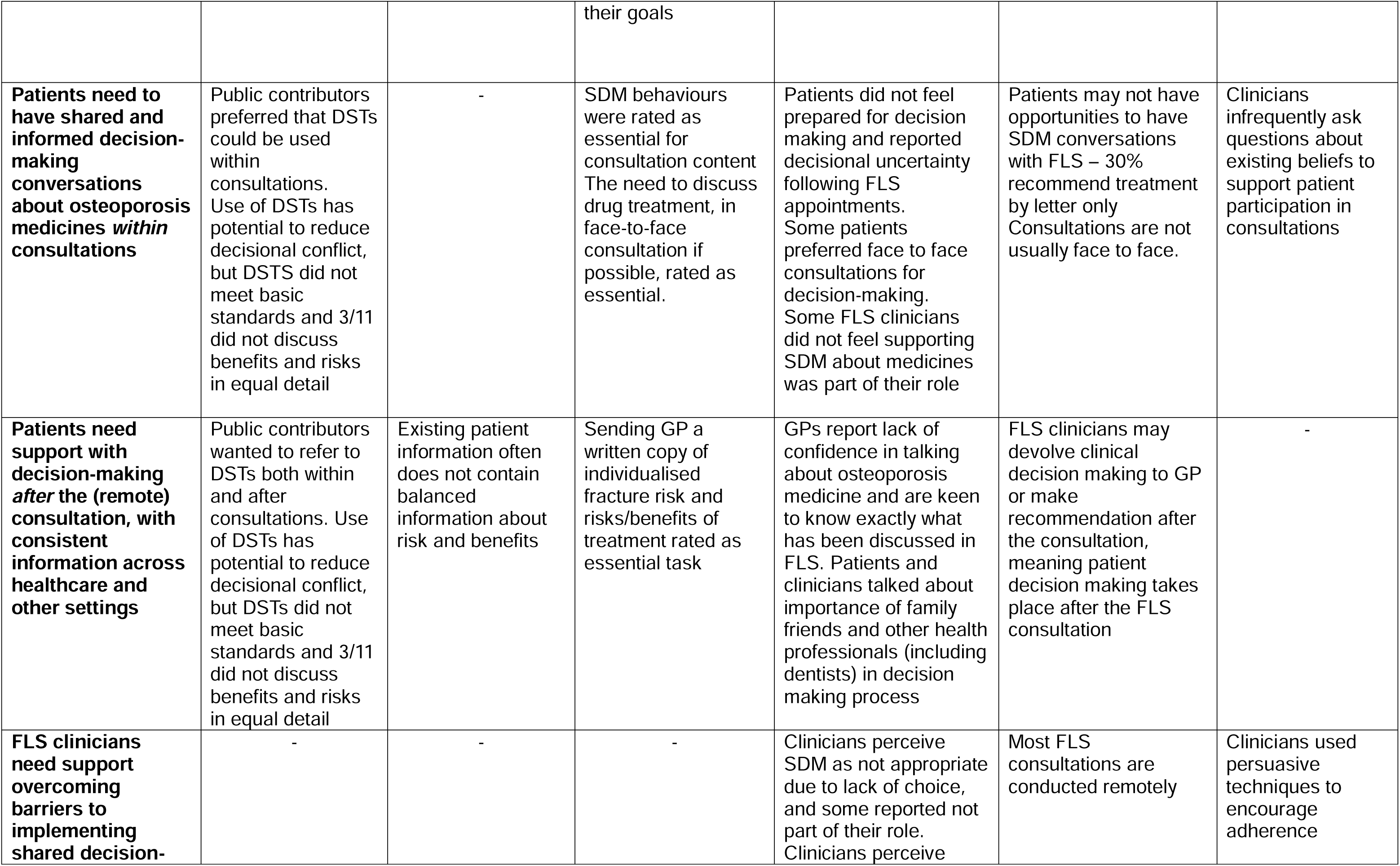
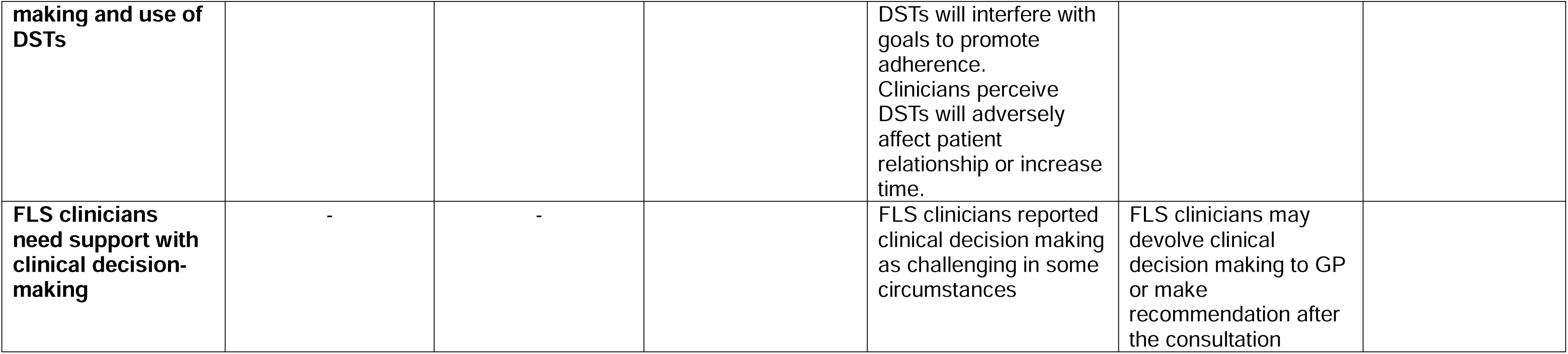
Joint display of key findings from iFraP development studies.

The qualitative findings[16] were used to identify a series of intervention design implication statements and questions, which were discussed within the team and the CoP.

An integration framework was developed, which brought together the underpinning theories and frameworks, the evidence gathered, universal precautions for health literacy, and public contributor and CoP discussions. This outlined the stages of the consultation and how either the DST or training would meet the needs identified.

#### Intervention design – DST

We employed an ‘informed’ design mode to make decisions about the prototype iFraP intervention design[39]. This means that we used CoP and public contributor input, alongside theory and evidence, as a conceptual framework to draft a storyboard outlining the DST’s key functions, structure, content, visuals and navigation (see Supplementary Figure S2).

Clinical drug recommendations and the benefits and risks of each osteoporosis medicine were underpinned by the best available scientific evidence and national clinical guidelines[40,41]. Clinical members of the iFraP team worked alongside expert advisors, including the chairs of the clinical guideline groups to develop the DST algorithm, in collaboration with the DST developer.

Before formal feasibility testing, the prototype DST underwent iterative internal testing cycles to identify bugs and review written and visual tool content. Clinicians ‘tested’ the tool’s algorithm using simulated patients. Clinicians and public contributors reviewed and provided feedback on usability, written and visual content.

#### Intervention design – clinician skills training

The iFraP Enhanced Consultation Skills Training Course was developed, integrating SDM theory and evidence alongside evidence-based behaviour change techniques (BCTs) to increase use of SDM consultation skills and the DST.

The barriers and enablers to behaviour change were identified using the TDF, in the qualitative intervention development study[16]. The TDF domains map to the Behaviour Change Wheel (BCW); the COM-B model of behaviour change which proposes three components for a given behaviour (‘B’) to occur: capacity (C), opportunity (O), and motivation (M)[42]. Understanding if the barriers to behaviour change are underpinned by ‘capacity’, ‘opportunity’ or ‘motivation’ guides the selection and incorporation of evidence-based strategies to facilitate behaviour change (known as ‘intervention functions’) and specific behaviour change techniques (BCTs), and the mode of delivery[42].

The APEASE criteria[42] helped the training development team (ZP,LB,JF,SR,JP) decide which of the appropriate BCTs should be integrated into the enhanced Consultation Skills Training Course, considering the affordability, practicability, (cost)effectiveness, acceptability, safety, and equity of the technique, in collaboration with the study team.

#### Intervention feasibility testing and refinement

Real-world testing of the prototype intervention was completed at one FLS site. Consenting FLS clinicians completed the Consultation Skills Training Course and completed three cycles of prototype testing with consenting FLS patients. Consultations were observed by a researcher and audio-recorded to explore intervention fidelity, using a pre-defined fidelity checklist.

Interviews were completed with each patient immediately after their consultation. After each cycle of testing, the FLS clinician(s) delivering the consultation was interviewed. Interviews were informed by topic guides, consultation observations and fidelity checklist findings.

Interviews were transcribed, and data were inductively coded, using a framework approach[33] by three qualitative researchers (LB,NT,MT), and then mapped to the TFA domains. Findings were discussed with the CoP to identify any required intervention refinements. More details of the feasibility testing procedures and methods are detailed in Supplementary Data S1.

#### Documenting the intervention

The intervention manual is integrated, in an interactive format, as part of the enhanced Consultation Skills Training Course with content detailing:

- intervention components and underpinning concepts
- what the intervention is hoping to achieve and why
- how the intervention was developed
- how to use the DST and integrate its use to suit patient needs and various clinical scenarios.

## Results

### Integration of key findings and intervention design

A summary of the meta-inferences or key findings from the development studies are shown in the joint display (Table 1). In brief, these meta-inferences demonstrate key ‘needs’ for the intervention to address:

1. Patients need an understanding of osteoporosis and/or its consequences, as important perceptual facilitators of adherence
2. Patients need information about osteoporosis and its treatment that is easily understandable
3. Patients need to have shared and informed decision-making conversations about osteoporosis medicines *within* consultations
4. Discussions about osteoporosis need to be personalised and person-centred
5. Patients need support with decision-making after the (remote) consultation, with consistent information across healthcare and other settings
6. FLS clinicians need support overcoming barriers to implementing SDM and use of DSTs
7. FLS clinicians need support with clinical decision-making

From the findings, key decisions were made relating to the scope and functionality of the intervention.

- A series of possible options for how the web-based DST could be used flexibly in telephone consults was considered by CoP and public contributors. Public contributors rejected the option to simultaneously view the DST with the clinician whist on the telephone, due to perceived cognitive burden. Instead, they agreed that the DST should be used by the clinician to guide the consultation and the patient receives a printout (personal ‘Bone Health Record’) after the consultation.
- Supporting information resources were important to address information needs after the consultation and consistency of messaging across healthcare providers and in other settings. Information resources include the DST printout (personal ‘Bone Health Record’) and a dentist card to explain osteoporosis medicines to dental care providers.
- Training modules identified related to SDM, health literacy and risk communication skills, but also a module outlining clear unambiguous language to talk about osteoporosis.

Components of the prototype intervention are described in Table 2. Table 3 details the relative roles of the DST and the training at different stages of the consultation, mapped to the underpinning theories and frameworks. Table 4 outlines the process of mapping the qualitative findings to the COM-B framework to identify suitable BCTs to include in the iFraP training course, addressing the identified barriers.

**Table 2:**
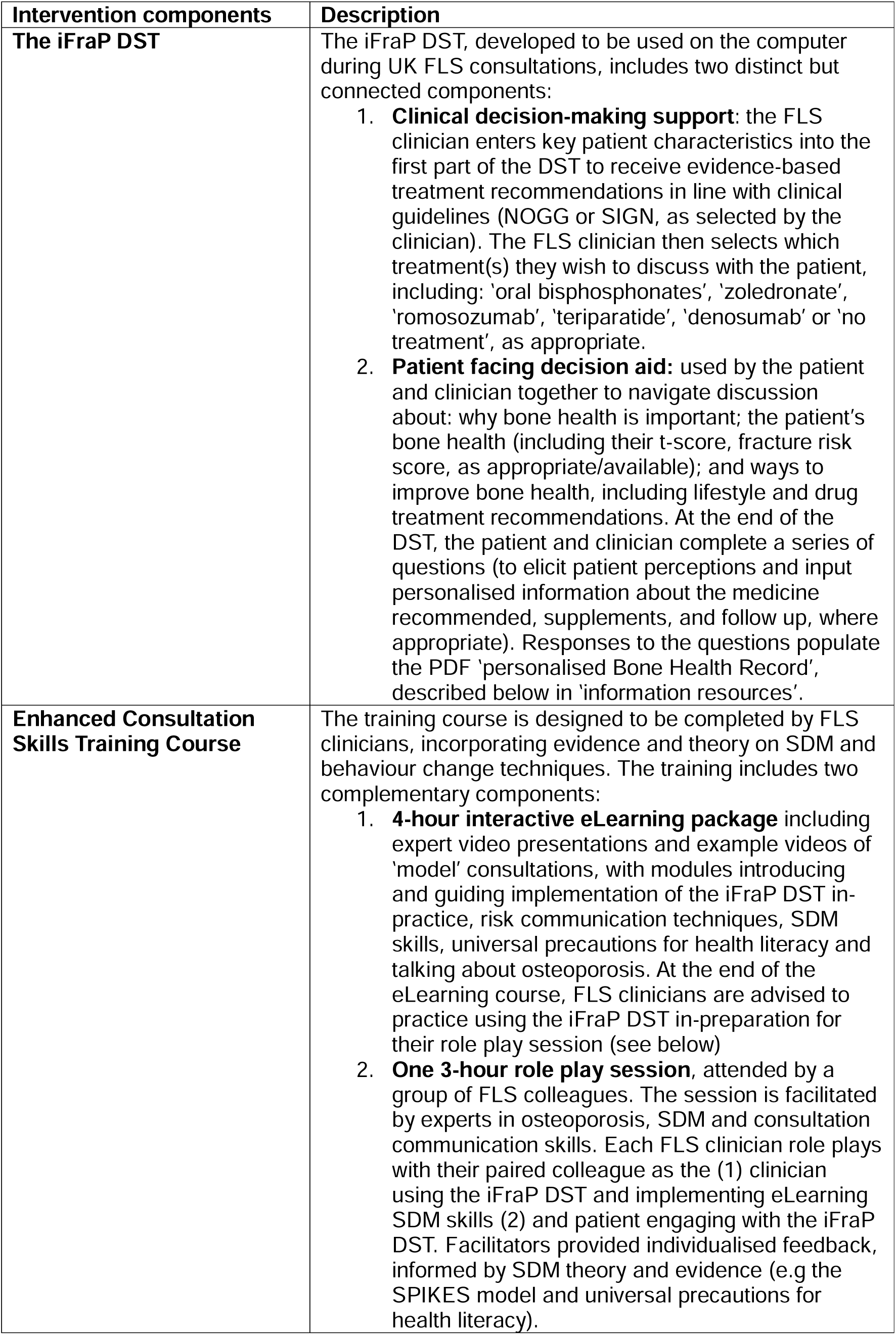
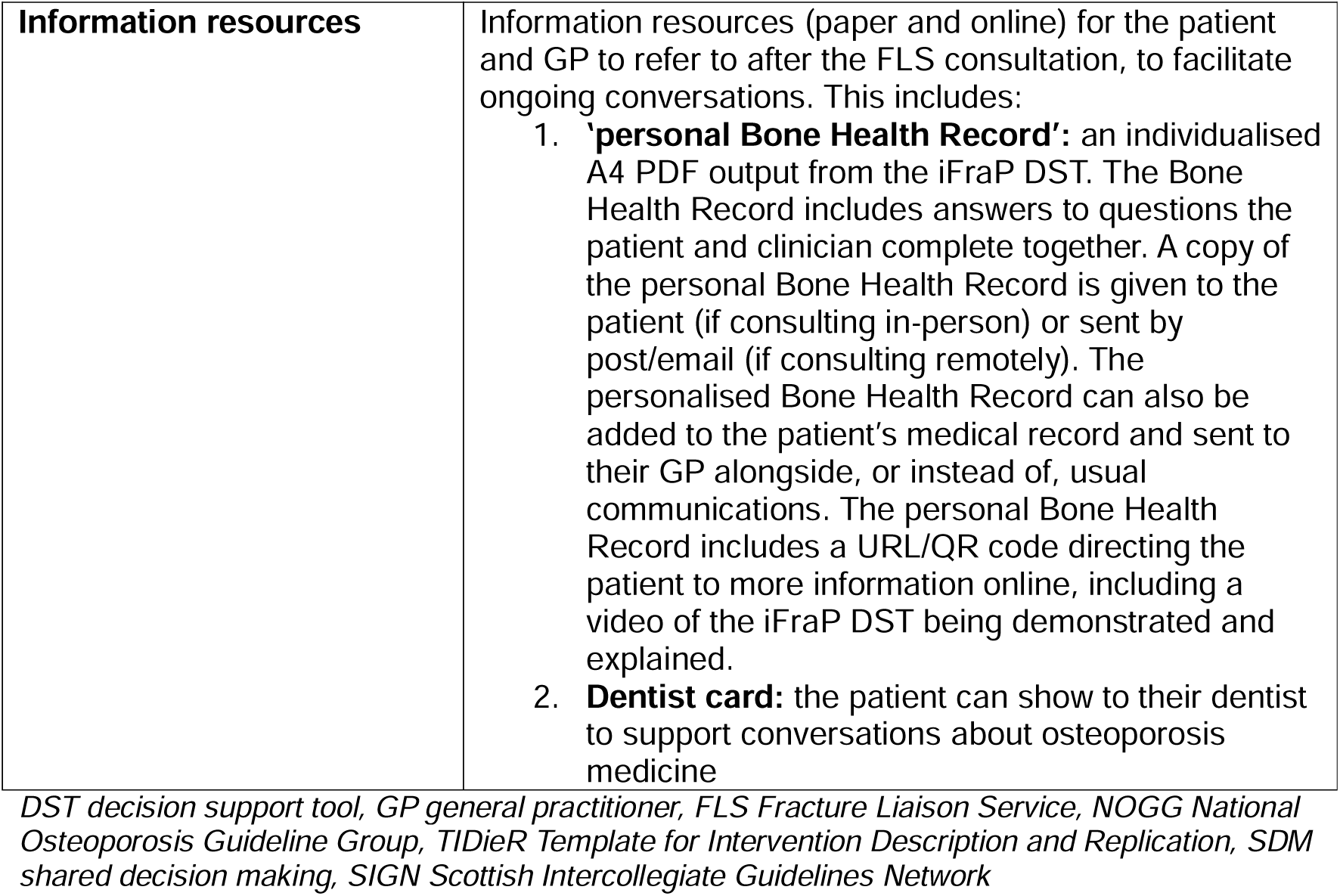
Final prototype iFraP intervention described using the TIDieR guidance.

**Table 3:**
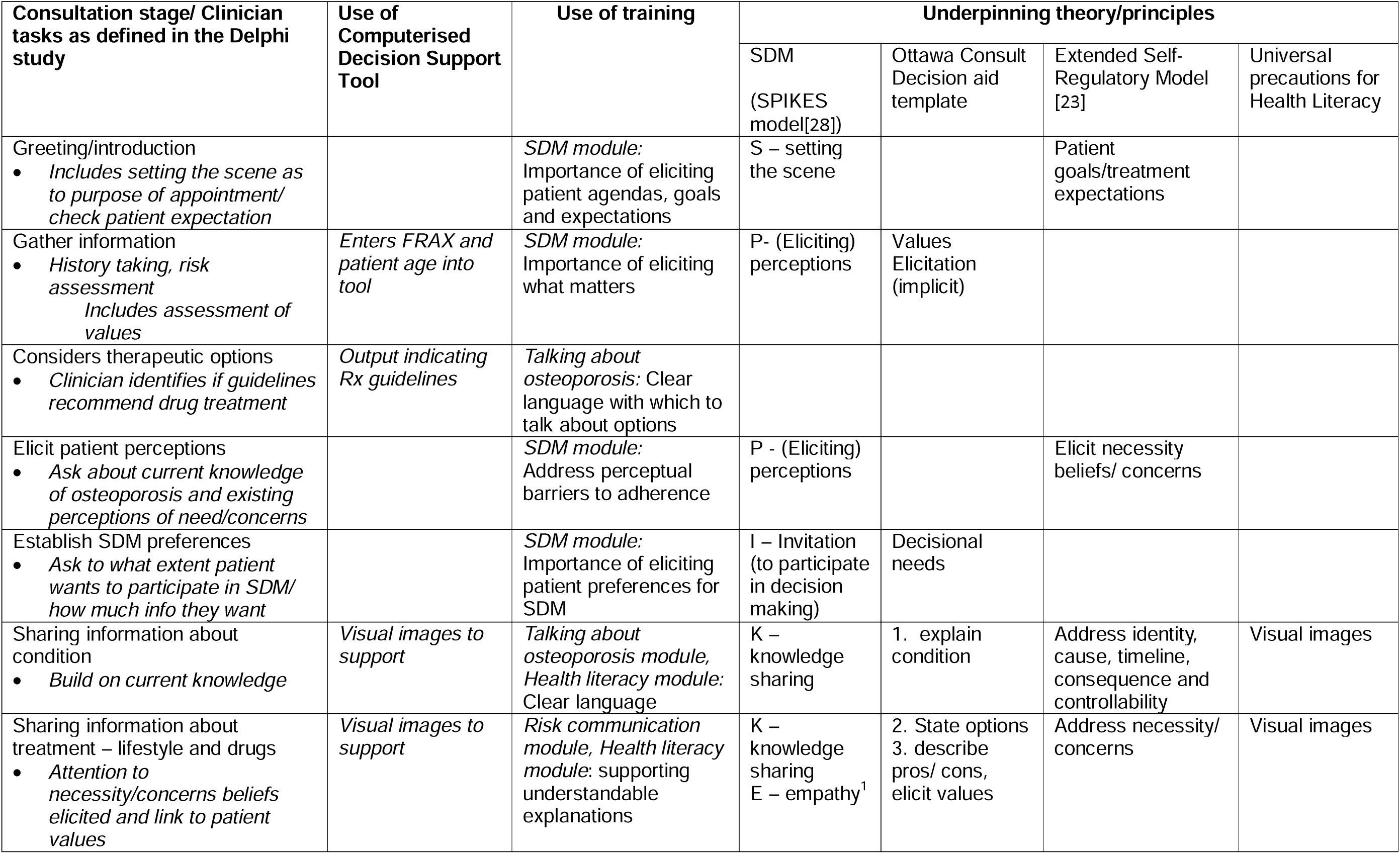
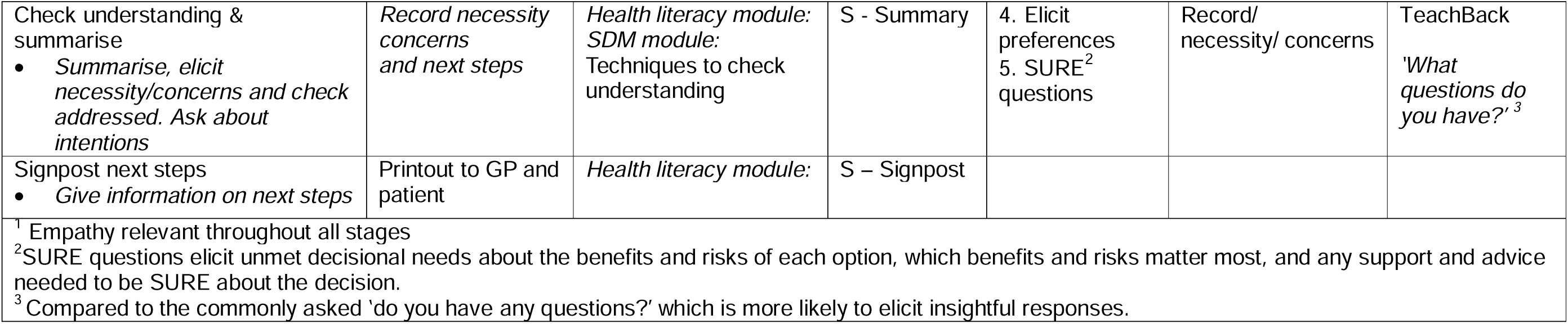
Consultation framework outlining role of DST and training, aligned to theories and frameworks.

**Table 4:**
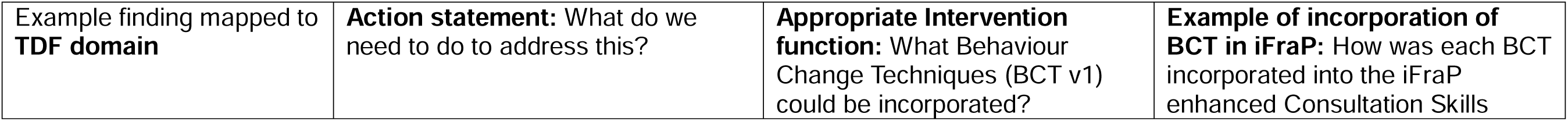
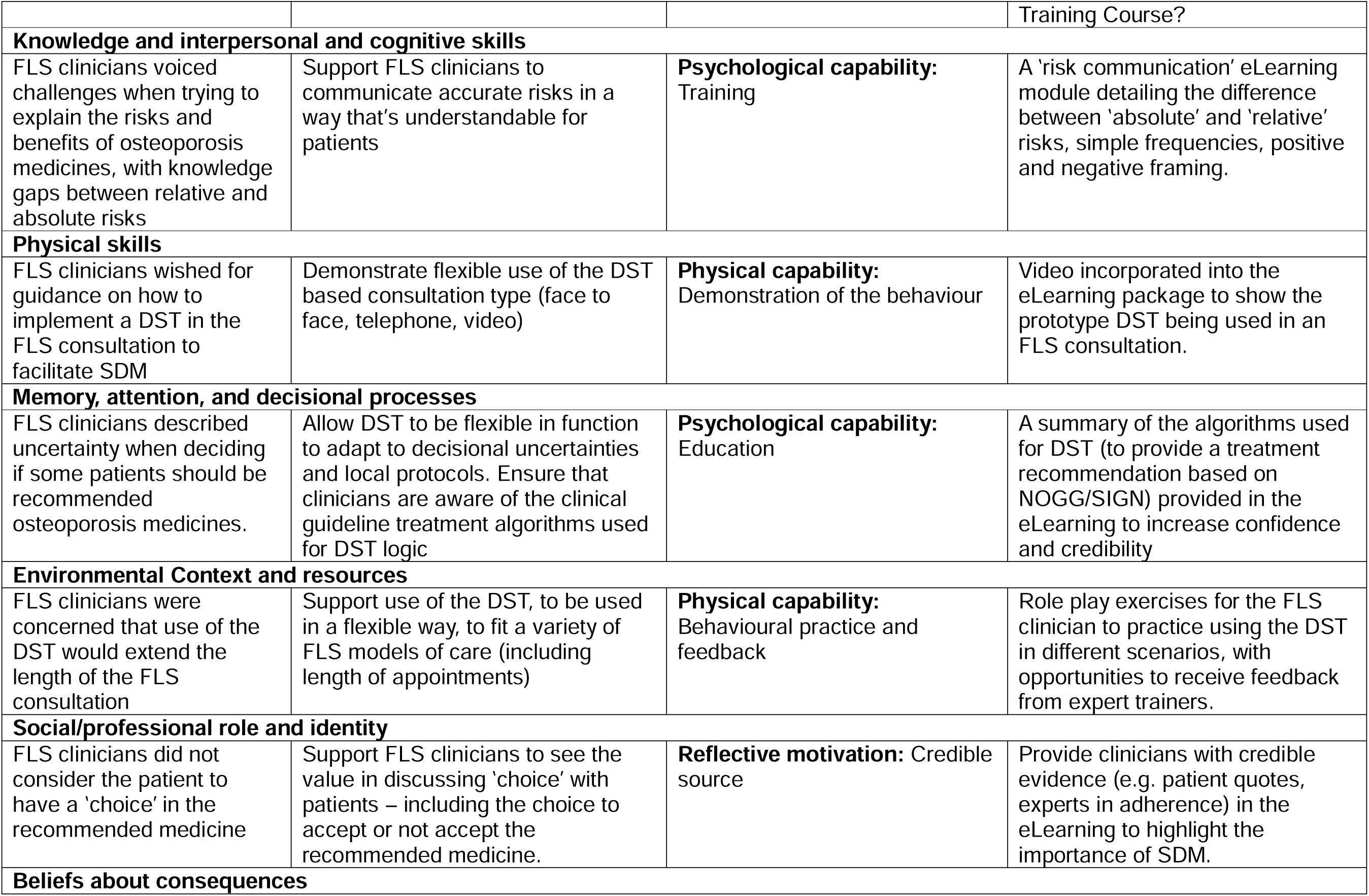
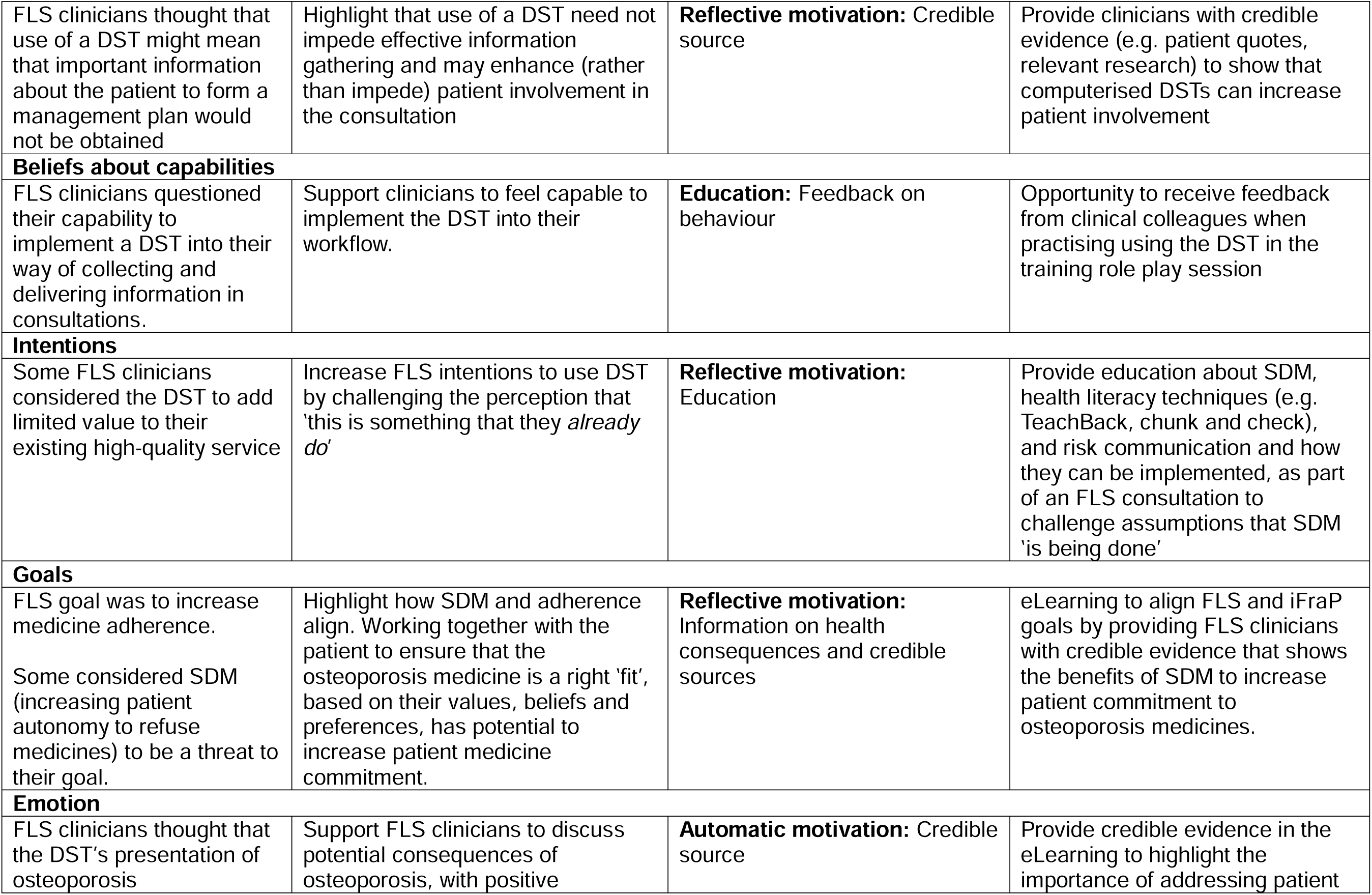
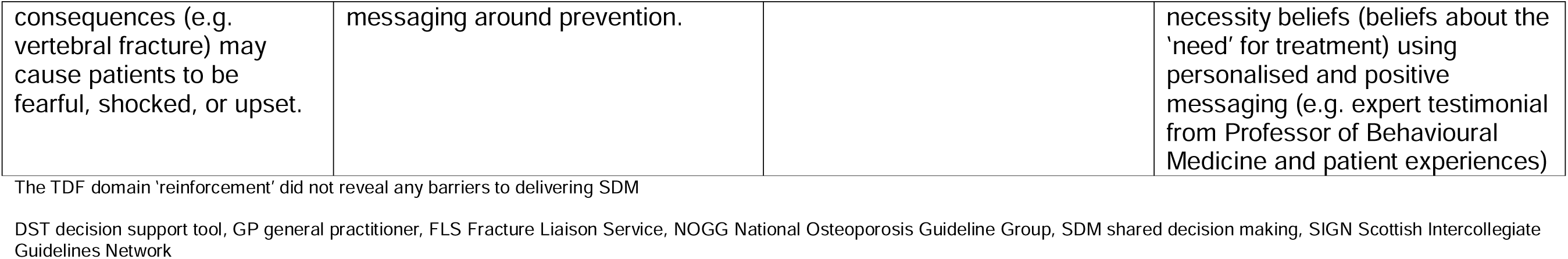
Use of the COM-B model to identify behaviour change techniques, mapped to the Theoretical Domains Framework domain.

### Intervention feasibility testing and refinement

Four clinicians working at Midlands Partnership University NHS Foundation Trust’s FLS in England completed the Enhanced Consultation Skills Training Course. FLS clinicians delivered iFraP consultations with 10 consenting FLS patients with a recent fragility fracture across three iterative testing cycles (n=3 cycle 1; n=3 cycle 2; n=4 cycle 3). Further details about participants are provided in Supplementary Data S1.

Overall, patients and FLS clinicians found the prototype iFraP intervention to be acceptable and feasible for use in FLS consultations. This was particularly evident through FLS clinicians’ and patients’ expressed wishes for iFraP to be used in future FLS appointments. iFraP was perceived as effective to achieve SDM by eliciting and addressing patient beliefs about osteoporosis and medicines, increasing patient involvement, and providing patients with sufficient and accessible information. However, FLS clinicians perceived that relaying information about the effectiveness of medicines and chance of side effects included in the DST may be a threat to adherence. FLS clinicians identified work (burden) required to implement iFraP, suggesting implementation could extend the consultation length. Increased allocated time to practice using the DST and opportunities to observe the DST ‘in-action’ were valued as ways to increase confidence and reduce overly structured delivery of the DST. Quotes, mapped to each TFA domain, as well as example updates made to the iFraP intervention, are presented in Table 5.

**Table 5.** Intervention feasibility testing findings, mapped the Theoretical Framework of Acceptability (TFA) domains.

| TFA Domain | Example quotes | Narrative interpretation | Example updates to the prototype iFraP intervention |
| --- | --- | --- | --- |
| **Affective attitude:** how patients and FLS clinicians feel about the iFraP intervention | “I thought it was fantastic (...) as well as looking at the screen she was explaining everything to you” (P07)<br>“You can actually see the diagrams, and like it’s on the screen, somebody’s like selecting it (...) it’s an opportunity [to say] ‘oh, can you just show me what this can cause if I did do this’. And as I say, that is just a brilliant idea being able to see it there” P09<br>“I think it’s all useful and relevant, I think it’s something I really will use time and time again. Definitely.” FLS04<br>“I don’t think for one second it isn’t a really valid good consultation and an improvement on probably what’s happening in fracture liaison services at the moment.” FLS01<br>“I feel like my consultations are better with it than they were without it and I do use it all the time (...) the bone health record I’m a massive fan of” FLS02 | Overall, patients and FLS clinicians liked the prototype iFraP DST, suggesting that the DST improved the quality of the consultation. | - |
| **Burden:** the perceived and experienced effort required to engage with, and deliver, the iFraP intervention | **Cognitive burden**<br>“It’s information overload isn’t it? You might be telling them why it’s important, but they haven’t had time to digest have they or think about why they’re taking it and stuff. So it’s just a bit soon to ask them.” FLS02<br>“I’m more thinking, ‘I’ll probably do it but I couldn’t 100% say because I need to go away and inwardly digest it.’ Everything tells me that I should be taking it. I think when you reflect back to the very first slide with the T-score, I could see that I was the lower end and that made it more sensible to take the medication.” P04 | Most patients described the content and structure of the iFraP DST as reducing cognitive burden.<br>On the other hand, some patients and FLS clinicians thought that using the DST might cause cognitive burden for patients by providing too much information that they require more time to digest before making a decision about medicines.<br>FLS clinicians also considered the burden of implementing the iFraP DST in clinical practice. Many reflected on the practise | FLS clinicians recommended altering the order of two components of the DST, allowing them to explain osteoporosis, before jumping into the patient’s results. This change was implemented to better align with the existing FLS workflow. |
|  | “It’s simple. You’re not bombarding somebody with something that they won’t understand. Very easy to understand” P09<br>**Work required to deliver iFraP in-clinic**<br>“I think it was a little bit tricky to start with because its, you know, different to what we are used to. But again, I think it’s something you just get used to. It did work, it worked for that patient, and she was, you know quite a challenging patient, it worked quite nicely, I thought, you know. So, its just getting used to it” FLS04<br>“there was nobody more sceptical about it at the beginning than me, I was thinking oh God here we go, another just extra work (...) I was really surprised at the timescale, I really thought that it would really add on to the clinic and put pressure on me. When I found out the timescale, it wasn’t really that much more and then in fact I think maybe it could really help a clinic run faster and more smoothly” FLS03<br>**Opportunity for iFraP to address service needs**<br>“there was some talk about us doing a report for the GPs and a report for the patient, so it’s two reports. We haven’t got time for that. So without too much work it addresses that. (...) I think the bone health assessment addresses the fact that we are sending the patient a report on what their bone density results are. To me it’s perfect in a very patient-friendly way (...) I just feel it addresses that gap that for our service we’ve been looking for and we haven’t been able to decide or to think what we could do and I feel that addresses it” FLS02 | required to integrate iFraP into their existing workflow and time limited FLS appointment. Some did consider how using the iFraP DST did not add pressures, and, in fact, the Bone Health Record posed opportunities to reduce work, by addressing current gaps in service provision. |  |
| **Ethicality:** the extent to which the iFraP intervention has | **Opportunity to be used with all patients**<br>“ideally, everyone should get it, shouldn’t they? I think it’s a better practice. I’m not putting anybody else down but it’s showing better care to people” P04 | Patients and FLS clinicians reflected that all patients should be given the opportunity to engage with the iFraP DST. One FLS clinician recommended that even patients | DST updated to include the option of ‘no treatment’, allowing the clinician to override the clinical recommendation produced by the |
| a good fit with the individual's value system | "Yes, I do think it's pretty fair and I do think that it should be made widely available" P01<br>"I think they are very fortunate aren't they to get it? You know it'd be great if all patients did get it and obviously that's the aim. It is unfair isn't it, in a way that some patients don't get it and others do." FLS04<br>"I think there should be an option that they should still have that, even if they don't have drug treatments, because you're still recommending things even though you're not recommending a treatment, you could be recommending lifestyle modifications, or you could be recommending that they up the calcium or..." FLS02<br>**Not appropriate for some groups**<br>"It depends how old your patient is as well. My patients can be elderly, so they're not really bothered about their bone health. All they want to do is give them a tablet and they'll quite happily go off and take it. Some people aren't. Some people are very engaged" FLS02 | who don't receive a drug recommendation should be given the opportunity to receive a Personalised Bone Health Record (produced by the DST).<br>Some FLS clinicians considered how some patient groups may have difficulty engaging with the DST, including those with sensory impairments. | DST algorithm. This allowed the Bone Health Record to be produced for people recommended 'no treatment' including scan results, lifestyle recommendations, and signposting.<br>Made DST text more visible, including increasing font size and changing font colours, where possible. |
| **Intervention coherence:** the extent to which the patient and FLS clinician understands the iFraP intervention and how it works | **Making sense of iFraP in FLS**<br>"He asked me questions and then supported it with things on the computer. It just felt more balanced and also there were no issues with answering questions because he answered the questions that I had. It just felt more of a conversation consultation rather than giving information which was quite nice." P02<br>"I think it's good and they should be doing this for quite a few things when you go and see a doctor or something. I think they should be speaking to you and showing you these things and explaining" P01<br>"For FLS services it's probably going to be invaluable (...), they're probably going to be very engaged in | **Making sense of iFraP in FLS**<br>Patients had a clear understanding of the purpose of the iFraP DST, reflecting that similar tools should be used in other clinical scenarios to facilitate shared discussions.<br>The iFraP training demonstrated to FLS clinicians the role of shared decision-making in medicine discussions, helping the intervention to make sense. After using the DST in practice, FLS clinicians could see how it aligned with the goals of their service and FLS more broadly. | Guidance to DST users that clinical judgement should prevail and its possible to override recommendations produced by the DST to abide to local protocols. |
|  | using it and wanting it" FLS02<br>"at first I was a bit sceptical and I thought well there's paper copies we do is pretty perfect and we've been doing that for years and I'm used to it, what's the point if it ain't broken, don't fix it. But I think after all this I think it's really good and I have enjoyed using it and I think it went really well" FLS03<br>**Alignment of iFraP to local protocols**<br>"because [the patient was] over 70 and it's based on the new NOGG [National Osteoporosis Guideline Group] it's like she could be treated because she'd had a fracture, but in actual fact we don't do that, it's over 75 (...) I don't agree with the over 70 either, I think that will cause a lot of controversy really."<br>FLS02 | **Alignment of iFraP to local protocols**<br>One clinician reflected that the clinical recommendation produced by the iFraP DST did not align with their local protocols. |  |
| **Opportunity costs:** the extent to which benefits, profits, or values must be given up to engage with, or deliver, the iFraP intervention | **Alignment of SDM and adherence goals**<br>“I find it awkward when I read side effects. You’ve sort of sold this idea of having this treatment because you want to prevent it and then you sort of say, ‘Now I’ll tell you about the side effects’ and kind of like putting them off it aren’t you?” FLS02<br>“we did the online training that research has shown that it’s quite important to find out what’s important to the patient and why it’s important to the patient to give help with them accepting the drug and staying on it. (...) the training did teach me it’s a shared agreement and if they don’t want treatment then, for me as a health professional, as long as I’ve provided them with a balanced view, it’s up to them what their decision is “FLS01<br>**Use of DST can feel procedural**<br>“I think it helps to see things visually, doesn't it? Perhaps it might feel a little bit - no, impersonal is the wrong word but they might feel it's a little bit impersonal.” P05<br>“she was focused on, I need to go through this tool and remember how it works. And which bits do I press here and whatever. (...) for me, it's not that I didn't find it personal, but it was very go through the motions of it.” P08<br>“So when I first started using the tool I was a little bit robotic with it.” IPT02 | **Alignment of SDM and adherence goals**<br>Medicine initiation and persistence are a Key Performance Indicators of FLS. Some FLS clinicians were concerned that elements of the iFraP DST would lead to a patient deciding not to start or continue taking an osteoporosis medicine. Engaging with the DST (and increasing patient autonomy to refuse medicines) may not align with the FLS's goal.<br>In contrast, one clinician reflected on the iFraP training, describing how shared decision-making and medicine adherence can be bedfellows.<br>**DST can feel procedural**<br>The structure provided by the DST was praised by patients and FLS clinicians to reduce patient’s cognitive burden (see ‘burden’) and guide FLS clinician information giving. However, some patients also reflected that the structured nature of the DST could lead to the consult feeling procedural, particularly when the DST was new or unfamiliar to the FLS clinician. FLS clinicians agreed, wanting more opportunities for greater familiarisation with the DST. | The iFraP eLearning was updated to include more guidance on practising using the DST with patients and observing the DST being used by other clinicians.<br>Increasing FLS clinician exposure to the DST was hoped to increase familiarisation with the tool’s content and function. |
| **Perceived (and experienced) effectiveness:** the extent to which the iFraP intervention was perceived, or experienced to | **Illness beliefs**<br>“it’s something I'd never even considered; that that’s what a bone looked like and that’s what happened. You just think it’s just something sort of white [laughter] and snaps like that, don't you? You don't think of the structure of a bone and the fact when it becomes osteopenic or osteoporotic, it’s actually thinner and then more likely to break. Again, it just | Patients and FLS clinicians described how the iFraP DST helped give understandable explanations of bone health and osteoporosis, the consequences of fracture, and the necessity and side effects of medicines, in a way that was personalised to the individual. |  |
| be, likely to achieve its purpose | makes it more understandable.” P04<br>“It’s like seeing the spines there and being able to visualise what your problem is. I think a visualisation always helps and it sinks in with what is going on with yourself.” P02<br>“when you see pictures like that with people bent over, I think it brings it home to you that you’ve got a health problem. You’ve just got to do the best you can for yourself haven’t you and live your life as you can like sort of thing.” P07<br>**Addressing necessity and concerns about medicines**<br>“The benefits, I think it again reconnects with the patient and why it’s important (...) that’s important to them and just finding out any issues that they may have and just hopefully so that they... so that it helps them to stay on the medication and if they’ve got any problems with it, you know, maybe that they can have alternative treatments so they just don’t stop taking it” FLS01<br>“she was worried about osteonecrosis of the jaw and I couldn’t remember the exact figures for it off the top of my head. I didn’t want to give her any false information so I quickly went through the tool (...) I do feel I’d addressed her concerns and she was still willing to go ahead” FLS02<br>“Obviously, there are side effects and I’d heard there was a lot of side effects on these Facebook pages that are supposed to be friendly and supportive. However, when she went through the numbers of how many people did get this and that, I found that was more beneficial and it kind of shot those people on Facebook out of the water that were saying not to take the tablets because of them” P02 | Most patients and FLS clinicians considered the DST to be effective at increasing patient involvement in the consultation. The structured DST and Personalised Bone Health Record were viewed as having potential to increase consistency of information and support ongoing conversations.<br>Patients and FLS clinicians considered how the iFraP intervention had potential to support patients to make decisions about medicines. |  |
|  | “He [FLS clinician] was going through it and being able to select the things that were important to me.” P09<br>**Patient involvement**<br>“He spoke to me as if like, I was a person. And not just a patient.” P09<br>“it felt more personal, more prompt, more, shall we say, modern and sophisticated” P08<br>“It involves them, it breaks the ice, you’re not going straight into, ‘oh you’ve got osteoporosis, you need this’. It kind of breaks the ice, gets them involved and gives you an idea of how the patient feels” FLS03<br>“It is more interactive with the patient, they do seem to get more involved with it” FLS01<br>**Providing understandable and consistent information**<br>“I think, you know its quite straight forward and easy for patients to understand” FLS04<br>“it’s all easy to understand. As far as I’m concerned, it’s all easy to understand.” P04<br>“the information was consistent. I said to [the patient], ‘It’s a new thing we’re doing’ and she said, ‘Oh yes, it was really good.’ She just emphasized she was really happy with the information and she was happy that the GP had got the same information” FLS02<br>“Everything went through in stages on it. It wasn’t just rushed, it was all the way through, different parts of it. And it kept you interested in what was going on.” P07 |  |  |
|  | “Going back to work, for instance (...) my boss was very concerned, so I think this will alleviate her concerns as well because I'll share [the Bone Health Record] with her.” P02<br>“I think it engages the patient to start with, when you say to them, you know ‘what do you understand?’ (...) I think is really useful because you know quite often they won’t be able to recall what you’ve said and then it just gives you that chance just to go over it again. so, I think that has worked well actually.” FLS04<br>**Decision-making about medicines**<br>“to see those three percentage points which is actually, you can improve by virtually a third, it puts it into perspective to say, yes, it is worth looking at here.” P08<br>“Everything tells me that I should be taking it. I think when you reflect back to the very first slide with the T-score, I could see that I was the lower end and that made it more sensible to take the medication. You go from the first slide to the last slide and it does make it a little bit more understandable as to why you should take it because you're that low in the score” P04<br>“by showing [the patient] the [iFraP DST] pictures and asking them extra questions, “how do you feel...” and involving her, I think that in my opinion swayed the patient to go on treatment. I think if it had just been a paper copy I think I'd have been struggling.” FLS03<br>**Use of iFraP in telephone consults**<br>“It's a visual aid for your patients and that isn't going to be the case with the telephone consultation. So I was apprehensive about using it, very sceptical that it<br>would work. But I was very pleasantly surprised actually. It was very helpful as more of probably a prompt to direct your consultation probably better than it would've been before." FLS02<br>"It depends how well she explains it because I can't see it, and she can't point 'it's this' or 'it's that'. If she says a particular type of part of the body or the actual bone density or anything, without seeing that picture how would I know?" P02<br>"Personally, no, because I'm not particularly computer literate [laughter]. I'm sure there are a lot of people who would find it difficult during a telephone consultation." P05 |  |  |
| **Self-efficacy:**<br>the patient's and FLS clinician's confidence that they can engage with, and deliver, the iFraP intervention | **Confidence delivering iFraP**<br>"I would be comfortable doing it as our standard way of doing our assessments" FLS01<br>"I felt confident because I'm experienced, I've been doing it 20 years. If you're going to catch me out on a question you know good luck to you. I felt confident using [the DST], everything on there I knew and I could use it and I could flirt around the screen" FLS03<br>"Quite confident really, it is quite straight forward. The little tabs on the side are nice and easy to navigate. I had a little practise beforehand." FLS04<br>"But saying that I don't know I'd react to a patient that said, oh no I don't want to take it and had a really low score where you go from there (...) the clinician needs maybe, I would like a little bit of training on what to do if the patient's if'ing and ah'ing or definitely doesn't want to take it and who does need it." FLS03<br>"Maybe just the more you do it the less awkward you | FLS clinicians overall reported feeling confident delivering the iFraP DST after completing the training skills course.<br>Some clinicians reflected that additional training would be valued to increase confidence delivering the DST in different scenarios (e.g. patients who do not want to take the medicine, telephone consultations)<br>Patients and FLS clinicians agreed that the DST increased confidence in the FLS's recommendations. | Additional videos were integrated into the eLearning. These videos demonstrated use of the DST and training skills in patient-clinician consultation scenarios, including:<br>1) Patients not recommended medicine<br>2) Telephone appointments<br>3) Comparing two medicine options<br>4) Patient not keen on taking a medicine<br>5) Referral to discuss intravenous medicines<br>6) Patient is already taking an osteoporosis medicine<br>The structure of the role play session was revised to allow clinicians increased allotted time to practise using the DST and communication skills with their colleagues and receiving feedback. |
|  | feel. As you settle into it you'll probably find your own little way of explaining it don't you, like anything, when anything's new." FLS02<br>"I recall having a lady last week who I was trying to use this format of the consultation and she started talking about how her fractures come from physical abuse from her husband and it's difficult because (...) I'm not trained really in and things that they may bring up around worry" FLS01<br>"more videos of different scenarios, that would have been useful. That's it really, I think it was good." FLS04<br>**iFraP increased confidence in FLS messaging**<br>"I think it's a good idea because they can see the information you're using and how they reach the points about yourself and your scores (...) I think seeing it for itself, people might just think, 'Yeah, maybe they are useful. Maybe they do know what they're talking about'" P02<br>"He asked me questions and then supported it with things on the computer." P04<br>"it wasn't just me talking it's there on the screen and it's like there's two of you here, if you don't believe me the computer doesn't lie kind of thing yeah, it's there in black and white on the computer. It isn't me just making it up" FLS03<br>"I think it probably helps just to reinforce that you know the treatment is necessary and that we've got the research and the tools behind that to back that up" FLS01 |  | Additional signposting added to the 'getting support' component of the DST, including a national domestic abuse helpline. These additions were also incorporated into the Bone Health Record. |

## Discussion

The iFraP intervention was developed through a rigorous, iterative and systematic approach, integrating existing evidence, frameworks and theories with primary data collection. Regular collaboration with stakeholders, expert advisors and public contributors ensured that intervention development centred on those who will deliver, use and benefit from it. Overall, the prototype iFraP intervention was perceived as acceptable and feasible to deliver in FLS, with potential to support SDM conversations about osteoporosis and related medicines.

Medicine adherence is optimised if a person believes it is necessary, relevant, safe, and practicable[43]. Concerns about side effects have been blamed for poor uptake of osteoporosis medicines, but together these studies show that patients are often unclear about the ‘need’ for osteoporosis medicines, particularly because osteoporosis itself is asymptomatic and misunderstood[24], contributing to difficulties making decisions about medicines[16].

Decision aids and DSTs have often been described as ‘requiring minimal training for use’. Consequently, few evaluations of DSTs have provided users with training to support implementation. In this development work, we identified the importance of complementary clinical and SDM training to overcome evidenced barriers to DST use[44,45]. At present, evidence demonstrating the potential for DSTs to improve medicine adherence outcomes is limited[4]. The feasibility study showed promise that the DST could support patient decision-making and uptake of medicines, underpinned by well-evidenced theories of medicine adherence[25,43]. Finally, our evidence synthesis of existing osteoporosis DSTs indicated that they were not ‘fit for purpose’ and rarely involved public contributors in their development[15]. The iFraP study integrated extensive public contribution throughout all aspects of development and testing, ensuring that it was understandable, relevant, and addressed their needs.

A key principle of intervention development is adapting to changing contextual factors to maximise intervention implementation in the real-world[12]. The iFraP study began in 2019, meaning that, intervention development required a flexible approach to continually assess and address uncertainties arising from a shifting context e.g. the move to remote consulting during and after the COVID pandemic, publication of NICE SDM guidelines[8], and introduction of new osteoporosis medicines into UK clinical practice[46]. In line with the MRC intervention development and testing guidance[10], we have sequentially separated ‘development’ and ‘feasibility testing’ in two distinct stages. However, the iterative and cyclical nature of development and feasibility testing allowed for continued refinement of the intervention to optimise relevance, acceptability and implementation.

### Limitations

Contextual understanding is essential when developing a complex intervention. The iFraP intervention was designed for use in UK FLS. Feasibility testing however was only conducted at one secondary care FLS site in England. Involvement of CoP stakeholders and expert clinical advisors increased the relevance of the intervention for use in Scotland (who follow the Scottish Intercollegiate Guidelines Network (SIGN) recommendations rather than the National Osteoporosis Guideline Group (NOGG) guidelines) and non-FLS care settings. However, we cannot assume transferability of the DST to FLS operating outside of the UK (where treatment guidelines may differ) or different practice settings (such as primary care). Further consideration would be required to determine how the intervention requires adaptation for use in non-FLS and non-UK contexts, and we have begun this process to adapt the resources for primary care use.

Four of the nine FLS clinicians who participated in the focus group study[16] also participated in the feasibility testing. This approach may have limited our understanding of varied FLS models of care and how the intervention might require adaptation to be implemented.

### Impact and Future Research

The iFraP development studies have already achieved impact. Our recommendations to improve the readability and quality of patient information about osteoporosis[13] led to national providers updating their information. Additionally, iFraP outputs are being incorporated into the ROS FLS implementation toolkit and findings have influenced updates to ROS Clinical Standards for FLS.

Our development work demonstrated while bone density scans are an essential component of FLS, where appropriate[17], information about bone density scans is often confusing and inconsistent[13]. These findings led to a successful funding application to explore how to optimise patient and clinician understanding of bone density scans and results[47].

The iFraP intervention will be tested in a pragmatic, parallel-group, individual RCT in four FLS sites in England (trial registration, ISRCTN55504164)[48], with nested mixed-methods process[49] and economic evaluations[50] to further test whether the intervention is effective, cost-effective, acceptable and implementable.

## Conclusion

We developed the iFraP intervention underpinned by evidence, theory, and extensive public contributor, patient, and clinician contribution. Feasibility testing demonstrated that the intervention was acceptable and feasible to use in UK FLS, with potential to improve patient outcomes. The iFraP RCT and nested economic and process evaluations will further evaluate implementation and effectiveness.

## Data Availability

All data produced in the present study are available upon reasonable request to the authors

## Ethics approval

Ethical permission for the iFraP development and feasibility studies was given by North West-Greater Manchester West Research Ethics Committee (reference number: 19/NW/0559).

## Acknowledgements

We would like to thank those that supported development of the iFraP intervention, including Dr Maddy Thompson, Dr Jane Fleming, Sarah Leyland, Professor Robert Horne, Professor Cynthia Paola Iglesias Urrutia, and Professor Sarah Ryan.

We would also like to acknowledge the important contributions of the Keele Osteoporosis Research User Group, the NIHR Clinician Scientist Award Steering Committee, the members of the study Keele Osteoporosis Community of Practice, and the participants themselves.

## Funding (specific to this study)

The iFraP intervention development and feasibility work was funded by the National Institute for Health and Care Research (CS-2018-18-ST2-010)/NIHR Academy], the Royal Osteoporosis Society, and Haywood Foundation. This study presents independent research funded by the NIHR. The views expressed are those of the author(s) and not necessarily those of the National Health Service (NHS), the NIHR, or the Department of Health and Social Care.

## References

1. Borgström F, Karlsson L, Ortsäter G, Norton N, Halbout P, Cooper C, et al. Fragility fractures in Europe: burden, management and opportunities. Arch Osteoporos [Internet] 2020 [cited 2021 Jun 23];15:59. Available from: https://link.springer.com/10.1007/s11657-020-0706-y

2. National Institute for Health and Care Excellence. Osteoporosis Products [Internet]. [cited 2025 Mar 21];Available from: https://www.nice.org.uk/guidance/conditions-and-diseases/diabetes-and-other-endocrinal--nutritional-and-metabolic-conditions/osteoporosis/products?ProductType=Guidance&Status=Published

3. Royal College of Physicians. You’ve had a fracture; how can we prevent another? Fracture Liaison Service Database (FLS-DB) [Internet]. 2025 [cited 2025 Mar 20]. Available from: https://www.rcp.ac.uk/media/ccllfgk1/fls-db-annual-report-2025.pdf

4. Stacey D, Légaré F, Lewis KB. Patient decision aids to engage adults in treatment or screening decisions [Internet]. JAMA - Journal of the American Medical Association2017 [cited 2020 Feb 21];318:657–8. Available from: http://jamanetwork.com/

5. Cornelissen D, de Kunder S, Si L, Reginster JY, Evers S, Boonen A, et al. Interventions to improve adherence to anti-osteoporosis medications: an updated systematic review. Osteoporosis International [Internet] 2020 [cited 2021 Dec 22];31:1645–69. Available from: https://link.springer.com/10.1007/s00198-020-05378-0

6. Kunneman M, Griffioen IPM, Labrie NHM, Kristiansen M, Montori VM, van Beusekom MM. Making care fit manifesto. BMJ Evid Based Med 2023;28:5–6.

7. NHS England. Universal personalised care: implementing the comprehensive model [Internet]. 2019 [cited 2025 Feb 11];Available from: https://www.england.nhs.uk/personalisedcare/comprehensive-model/

8. National Institute for Health and Care Excellence. Shared decision making [Internet]. [cited 2020 Jan 21];Available from: https://www.nice.org.uk/about/what-we-do/our-programmes/nice-guidance/nice-guidelines/shared-decision-making

9. Paskins Z, Jinks C, Mahmood W, Jayakumar P, Sangan CB, Belcher J, et al. Public priorities for osteoporosis and fracture research: results from a general population survey. Arch Osteoporos [Internet] 2017 [cited 2019 Dec 23];12:45. Available from: http://link.springer.com/10.1007/s11657-017-0340-5

10. Skivington K, Matthews L, Simpson SA, Craig P, Baird J, Blazeby JM, et al. A new framework for developing and evaluating complex interventions: update of Medical Research Council guidance. BMJ [Internet] 2021 [cited 2021 Oct 4];374:n2061. Available from: 10.1136/bmj.n2061

11. Grol R, Wensing M, Eccles M. Improving patient care: The implementation of change in clinical practice. Edinburgh: Butterworth Heinemann.; 2005.

12. O’Cathain A, Croot L, Duncan E, Rousseau N, Sworn K, Turner KM, et al. Guidance on how to develop complex interventions to improve health and healthcare. BMJ Open 2019;9:e029954.

13. Crawford-Manning F, Greenall C, Hawarden A, Bullock L, Leyland S, Jinks C, et al. Evaluation of quality and readability of online patient information on osteoporosis and osteoporosis drug treatment and recommendations for improvement. Osteoporosis International [Internet] 2021 [cited 2021 Aug 23];32:1567–84. Available from: https://link.springer.com/article/10.1007/s00198-020-05800-7

14. Bullock L, Abdelmagid S, Fleming J, Leyland S, Clark EM, Gidlow C, et al. Variation in UK Fracture Liaison Service consultation conduct and content before and during the COVID pandemic: results from the iFraP-D UK survey. Arch Osteoporos 2023;

15. Paskins Z, Torres-Roldan V, Hawarden A, Bullock L, Urtecho LM, Torres G, et al. Quality and effectiveness of osteoporosis treatment decision aids: a systematic review and environmental scan. Osteoporosis International 2020;

16. Bullock L, Manning F, Hawarden A, Fleming J, Leyland S, Clark EM, et al. Exploring practice and perspectives on shared decision-making about osteoporosis medicines in Fracture Liaison Services: the iFraP development qualitative study. Arch Osteoporos 2024;19:50.

17. Bullock L, Crawford-Manning F, Cottrell E, Fleming J, Leyland S, Edwards J, et al. Developing a model Fracture Liaison Service consultation with patients, carers and clinicians: a Delphi survey to inform content of the iFraP complex consultation intervention. Arch Osteoporos [Internet] 2021 [cited 2021 Aug 5];16:1–17. Available from: https://link.springer.com/article/10.1007/s11657-021-00913-w

18. Paskins Z, Bullock L, Crawford-Manning F, Cottrell E, Fleming J, Leyland S, et al. Improving uptake of Fracture Prevention drug treatments: a protocol for Development of a consultation intervention (iFraP-D). BMJ Open [Internet] 2021 [cited 2021 Aug 19];11:e048811. Available from: https://bmjopen.bmj.com/content/11/8/e048811

19. Duncan E, O’Cathain A, Rousseau N, Croot L, Sworn K, Turner KM, et al. Guidance for reporting intervention development studies in health research (GUIDED): an evidence-based consensus study. BMJ Open 2020;10:e033516.

20. McLellan AR, Gallacher SJ, Fraser M, McQuillian C. The fracture liaison service: success of a program for the evaluation and management of patients with osteoporotic fracture. Osteoporosis International 2003;14:1028–34.

21. Wenger E, McDermott RA, Snyder W. Cultivating communities of practice: A guide to managing knowledge. Boston, Mass: Harvard Business School Press.2002;

22. Horne R, Weinman J. Self-regulation and Self-management in Asthma: Exploring The Role of Illness Perceptions and Treatment Beliefs in Explaining Non-adherence to Preventer Medication. Psychol Health 2002;17:17–32.

23. Leventhal H, Benyamini Y, Brownlee S, Diefenbach M, Leventhal EA, Patrick-Miller L, et al. Illness representations: Theoretical foundation. In: Perceptions of health and illness: Current research and application. Amsterdam: Harwood Academic Publisher; 1997.

24. Barker KL, Toye F, Lowe CJM. A qualitative systematic review of patients’ experience of osteoporosis using meta-ethnography. Arch Osteoporos 2016;11:33.

25. Horne R, Chapman SCE, Parham R, Freemantle N, Forbes A, Cooper V. Understanding Patients’ Adherence-Related Beliefs about Medicines Prescribed for Long-Term Conditions: A Meta-Analytic Review of the Necessity-Concerns Framework. PLoS One [Internet] 2013;8:e80633. Available from: https://dx.plos.org/10.1371/journal.pone.0080633

26. NICE. Medicines adherence: involving patients in decisions about prescribed medicines and supporting adherence [Internet]. NICE; 2009 [cited 2020 Jul 24]. Available from: https://www.nice.org.uk/guidance/cg76/chapter/1-Guidance#patient-involvement-in-decisions-about-medicines

27. Stacey D, O’Conner AM. Ottawa Consult Decision Aid template [Internet]. In: O’Connor A, Stacey D, Jacobsen M, editors. Ottawa Decision Support Tutorial (ODST): Improving Practitioners’ Decision Support Skills Ottawa Hospital Research Institute: Patient Decision Aids. 2011. Available from: https://decisionaid.ohri.ca/ODST/

28. Baile WF, Buckman R, Lenzi R, Glober G, Beale EA, Kudelka AP. SPIKES—A Six-Step Protocol for Delivering Bad News: Application to the Patient with Cancer. Oncologist [Internet] 2000 [cited 2020 Oct 16];5:302–11. Available from: https://pubmed.ncbi.nlm.nih.gov/10964998/

29. McCaffery KJ, Holmes-Rovner M, Smith SK, Rovner D, Nutbeam D, Clayman ML, et al. Addressing health literacy in patient decision aids. BMC Med Inform Decis Mak [Internet] 2013 [cited 2020 Jul 14];13:S10. Available from: https://bmcmedinformdecismak.biomedcentral.com/articles/10.1186/1472-6947-13-S2-S10

30. NHS Scotland. The Health Literacy Place: Techniques.

31. Naik G, Ahmed H, Edwards AG. Communicating risk to patients and the public. British Journal of General Practice 2012;62:213–6.

32. Talevski J, Wong Shee A, Rasmussen B, Kemp G, Beauchamp A. Teach-back: A systematic review of implementation and impacts. PLoS One [Internet] 2020 [cited 2021 Feb 24];15:e0231350. Available from: https://dx.plos.org/10.1371/journal.pone.0231350

33. Gale NK, Heath G, Cameron E, Rashid S, Redwood S. Using the framework method for the analysis of qualitative data in multi-disciplinary health research [Internet]. 2013 [cited 2020 Jan 14]. Available from: http://www.biomedcentral.com/1471-2288/13/117

34. Cane J, Connor DO, Michie S. Validation of the theoretical domains framework for use in behaviour change and implementation research. 2012;1–17.

35. Sekhon M, Cartwright M, Francis JJ. Acceptability of healthcare interventions: An overview of reviews and development of a theoretical framework. BMC Health Serv Res [Internet] 2017;17:1–13. Available from: 10.1186/s12913-017-2031-8

36. Hawarden AW, Bullock L, León-García M, Hartasanchez SA, Hargraves I, Horne R, et al. P083 How are beliefs about osteoporosis medicines elicited and addressed in clinical consultations: a video observational study. Rheumatology 2023;62.

37. Paskins Z, Bullock L, Manning F, Bishop S, Campbell P, Cottrell E, et al. Acceptability of, and preferences for, remote consulting during COVID-19 among older patients with two common long-term musculoskeletal conditions: findings from three qualitative studies and recommendations for practice. BMC Musculoskelet Disord [Internet] 2022;23:312. Available from: https://bmcmusculoskeletdisord.biomedcentral.com/articles/10.1186/s12891-022-05273-1

38. Paskins Z, Crawford-Manning F, Bullock L, Jinks C. Identifying and managing osteoporosis before and after COVID-19: rise of the remote consultation? Osteoporosis International [Internet] 2020 [cited 2020 Oct 16];31:1629–32. Available from: 10.1007/s00198-020-05465-2

39. Rousseau N, Turner KM, Duncan E, O’Cathain A, Croot L, Yardley L, et al. Attending to design when developing complex health interventions: A qualitative interview study with intervention developers and associated stakeholders. PLoS One 2019;14:1–20.

40. National Osteoporosis Guideline Group. NOGG 2021: Clinical guideline for the prevention and treatment of osteoporosis. 2021.

41. Scottish Intercollegiate Guidelines Network. Management of osteoporosis and the prevention of fragility fractures: A national clinical guideline [Internet]. 2015 [cited 2020 Oct 19]. Available from: www.sign.ac.uk/guidelines/fulltext/50/index.html

42. Michie S, Atkins L, West R. The Behaviour Change Wheel: A Guide to Designing Interventions. London: Silverback Publishing; 2014.

43. National Institute for Health and Care Excellence. Medicines adherence: involving patients in decisions about prescribed medicines and supporting adherence . 2009;

44. Joseph-Williams N, Lloyd A, Edwards A, Stobbart L, Tomson D, Macphail S, et al. Implementing shared decision making in the NHS: lessons from the MAGIC programme. BMJ [Internet] 2017 [cited 2020 Jul 25];357:j1744. Available from: http://www.bmj.com/permissionsSubscribe:http://www.bmj.com/subscribeBMJ2017;357:j17 44doi:10.1136/bmj.j1744

45. Joseph-Williams N, Abhyankar P, Boland L, Bravo P, Brenner AT, Brodney S, et al. What Works in Implementing Patient Decision Aids in Routine Clinical Settings? A Rapid Realist Review and Update from the International Patient Decision Aid Standards Collaboration. Med Decis Making [Internet] 2020 [cited 2021 Apr 8];272989X20978208. Available from: http://www.ncbi.nlm.nih.gov/pubmed/33319621

46. National Institute for Health and Care Excellence. Romosozumab for treating severe osteoporosis. 2022

47. Kettle C, Butterworth J, Griffin J, Henderson B, Jinks C, Knapp K, et al. P749 Exploring patient and clinician understanding of bone density (DXA) scan results: a qualitative study. Aging Clin Exp Res 2024;36:174.

48. Bullock L, Nicholls E, Cherrington A, Butler-Walley S, Clark EM, Fleming J, et al. A person-centred consultation intervention to improve shared decision-making about, and uptake of, osteoporosis medicines (iFraP): a pragmatic, parallel-group, individual randomised controlled trial protocol. NIHR Open Research 2024;4:14.

49. Bullock L, Cherrington A, Clark EM, Fleming J, Bentley I, Nicholls E, et al. Protocol for a mixed methods process evaluation for a randomised controlled trial to improve shared decision-making about, and uptake of, osteoporosis medicines: the iFraP study. NIHR Open Research 2024;4:70.

50. Siciliano M, Bathers S, Bentley I, Bullock L, Cherrington A, Clark E, et al. Protocol for a trial-based economic evaluation analysis of a complex digital health intervention including a computerised decision support tool: the iFraP intervention. NIHR Open Research 2024;4:15.

