## Supplementary Table S1 for "The Michael Mason Prize: Development and feasibility testing of a complex intervention to improve adherence to fracture prevention medicine using a person-centred approach"

### Supplementary Table S1: Public contributor ‘impact log’, presented chronologically through the development and testing of iFraP

| **Before iFraP development** | | | |
| --- | --- | --- | --- |
| **1 PPIE activity** | **2 Activity objectives** | **3 Take homes from PPIE activity** | **4 Impact on development / trial / other –**  **What changes were made?** |
| Setting the context for iFraP: pre-iFraP PPIE work | To discuss focus group data^1^ to understand priorities for osteoporosis and fracture research | 1. Public contributors identified ‘information needs’ as an important (yet missing) theme in the analysis of focus group data^1^. The theme related to unmet ‘information needs’ went onto be included in a national survey priority setting exercise^2^. In this survey, ‘improving information from health professionals’ rated as the highest-rated research priority | National survey^2^ demonstrated that improving information from health professionals was the highest-rated research priority, underpinning the idea for iFraP. |
| **iFraP Development and Feasibility testing (iFraP-D)** | | | |
| **1 PPIE activity details** | **2 Activity objectives** | **3 Take homes from PPIE activity** | **4 Impact on development / trial / other –**  **What changes were made?** |
| Public Contributor meeting 1  (n=5) | To discuss patient facing documents to be submitted to ethics for iFraP development work | 1. Recommendations to update documents to make information for understandable and accessible such as removal of ‘iFraP intervention’ throughout, instead focusing on the aims of the intervention ‘to improve the way nurses, doctors and clinicians communicate with patients about osteoporosis’ | 1. Study documents updated before submission to ethics committee |
| Community of Practice meeting 1  (n=2 public contributors, Total of 10 attendees) | To bring together different stakeholders to discuss thoughts about iFraP | Public contributors and stakeholders told us:   1. ‘Fractures’ is often misinterpreted by patients 2. The consequences of osteoporotic fractures are not clear and need to be personalised to what ‘the person wants to do in their lives’ 3. Osteoporosis is often seen as a ‘woman’s disease’ | 1. Use ‘broken bones’ instead of fractures 2. Need to emphasise the potential of the biopsychosocial impacts of osteoporosis, tailored to the individual 3. Be inclusive to ensure osteoporosis is seen as a condition that affects men and women |
| Community of Practice meeting 2  (n=2 public contributors, Total of 11 attendees) | To discuss the purpose and function of the DST in FLS and discuss optimal language for describing osteoporosis, using evidence from patient information resources review | Public contributors and stakeholders told us:   1. To think about how we support the patient to continue to have informed discussions with GPs/friends/family/dentist after the consultation 2. That terms such as ‘spongy’, ‘fragile’ to describe bones induces fear 3. That terms such as ‘prevent’ ‘renew’ and ‘restore’ are misleading when describing the actions of osteoporosis medicines | 1. iFraP information resources (Personalised Bone Health Record and dentist card) to meet needs 2. Use ‘less strong’ or ‘weaker’ to describe bone strength 3. Use ‘strengthening bone’ and ‘lowering the chance of future fracture’ when describing the benefits of osteoporosis medicines |
| Public Contributor meeting 2 | To discuss iFraP Delphi survey content and format on Health Survey | 1. Noted that survey wasn’t representative 2. Recommended updates in wording and presentation of the survey for ease of completion and sense | 1. Changes to make survey to make more representative: allow participants to enter younger age groups, vignettes to include male and female examples (rather than just female) 2. Recommended edits made, including: separating single statements into multiple, providing examples, such as: ‘ask the patient about their goals in life, *eg continuing to live independently, travel the world’* to increase ease of completion |
| Pilot the Delphi survey  (n=2) | To pilot Delphi survey with public contributors for feedback | 1. Provided summary of how long survey took to complete and provided recommended updates | 1. Updates made to Delphi survey based on public contributor feedback, including typos and sense |
| Two Delphi survey analysis meetings, each including two public contributors | To gain public contributor insights and data interpretations of Delphi rounds 1 and 2 | 1. Public contributors considered if, based on the findings, survey statements should be removed or be amended. If amended, public contributors helped to reword statements. | 1. Public contributors directly supported the inclusion of statements that informed recommendations for a model FLS consultation (produced by the Delphi survey) |
| Public Contributor meeting 3  (n=2) | To discuss focus group topic guides to ensure they are understandable and covered priority areas | Public contributors told us that:   1. ‘Involvement’ was a priority area to explore – the importance of a two-way conversation 2. ‘Appointment’ was more understandable than ‘consultation’ 3. Talking about a decision aid and ‘fracture risk’ in the focus group would not be clear. Hard to visualise a decision aid, and lots of medical concepts such as ‘risk’ are abstract and might limit the quality of data collected | 1. Updated topic guide to ask questions about patient involvement in the discussion 2. Updated wording in patient facing materials (appointment rather than consultation) 3. Showed images as part of the focus groups, for example, a ‘mock up’ of a decision aid and risk Cates Plot to facilitate discussions |
| Public contributor meeting 4  (n=4) | To discuss the patient focus group data and explore data interpretations | 1. Discussed the concept of ‘choice’ - Important that ‘alternative’ medicine choices are given with contextual information i.e. when they would be offered, and under what circumstances (such as in the event of gastrointestinal side effects) 2. Should present benefits and risks against each other to show a balanced view 3. Importance written materials to reduce the cognitive load of receiving a diagnosis and making a decision about medicines | 1. Added to the clinician training: important for clinicians to explain that alternative options are available and in what circumstances 2. Added to DST: Cates plots that include benefits and risks together for comparison 3. Added personalised Bone Health Record: to complement current FLS materials to provide accessible, understandable and consistent information |
| Community of Practice meeting 3  (n=2 public contributors, 1 Royal Osteoporosis Society patient advocate. Total of 10 attendees) | To discuss use of the iFraP computerised decision support tool in remote consultations (COVID) | Public contributors and Royal Osteoporosis Society patient advocates told us:   1. Use of the iFraP DST during the consultation must be flexible to accommodate for patient’s ability to use technology. Consider providing patients with information before and after the consultation and use of the tool remotely, if possible. | 1. Essential to design iFraP flexibly to be used in telephone consultations because of service changes during COVID. |
| Video stars for iFraP prototype intervention  (n=2) | To support the development of videos to be integrated into the iFraP eLearning training | 1. Public contributors supported development of iFraP prototype eLearning videos    a. Patient testimonial of the importance of being involved in discussions about osteoporosis medicines    b. Public contributor role played as ‘patient’ in mock consultation using the iFraP prototype DST | 1. Supported the development of the iFraP eLearning in preparation for the feasibility testing study. Feedback from FLS clinicians that these videos were an essential feature of the eLearning course. |
| Community of Practice meeting 4  (n=1 public contributor, 1 Royal Osteoporosis Society patient advocate. Total of 12 attendees) | To discuss content (pictures, text and links) of the iFraP DST | Public contributors and Royal Osteoporosis Society patient advocates told us:   1. ‘Osteopenia’ can be an unhelpful term for older adults 2. T-scores are difficult to understand and graphics/images that show these are helpful 3. To demonstrate the impact of spinal fractures, using images, as these are often not recognised as a potential consequence of osteoporosis | 1. Preference for ‘low bone density’ rather than ‘osteopenia’. 2. Inclusion of t-score imagery to be included in the DST and Personalised Bone Health Record 3. Inclusion of an animation to demonstrate the impact of spinal fractures |
| Public contributor meeting 5  (n=5) | To discuss the design of the DST, including content, images, and function | Public contributors told us:   1. Images of skeleton were synonymous with ‘death’ 2. the importance of recognizing both males and females when presenting risk factors of osteoporosis – rather than focusing on the menopause 3. When discussing consequences of a hip fracture they preferred a wheelchair image rather than a hospital bed to emphasise reliance on others (rather than short hospital stay which wouldn’t impact their lives long-term) 4. When presenting ‘Keeping active’ – the imagery should represent exercise and broader physical ‘activities’ e.g. aerobics, walking 5. Terms such as ‘osteonecrosis of the jaw’ and Atypical Femur Fracture are not understandable. 6. Happy with simple frequencies (e.g. 1/1000). Wished for terms such as ‘rare’ and ‘common’ to complement simple frequencies for people that aren’t as confident with numbers. | 1. Alternative skeleton images chosen, including an outline of a body to represent the living skeleton. 2. Tool updated to say ‘low sex hormones’ as a risk factor rather than ‘menopause’ 3. Inclusion of a wheelchair image to represent the biopsychosocial impact of hip fracture 4. Inclusion of yoga image rather than ‘gym’ image to overcome barriers to inclusion 5. Public contributors agreed use of terms: ‘jawbone problem’ (ONJ) and ‘unusual thigh bone break’ (AFF) 6. Inclusion of words ‘common’ and ‘very rare’ as different people with prefer different ways of communicating risk |
| Public contributor meeting 6  (n=3) | To discuss the DST design, delivery (in remote consultations) and  iFraP feasibility testing research design | Public contributors told us:   1. They were concerned too much information would be provided on the tool if there were details about the uncertainty of evidence and where the evidence has come from. 2. When discussing uncertainty about the evidence (e.g. 1 in 10,000 or 1 in 100) all preferred ‘approx’ rather than ‘about’ 3. Preferred not to receive access to the tool during a telephone consultation. This would cause cognitive overload and difficulties with navigation. All public contributors preferred the printout sent to them after the consultation. This was reported back to the Trial Steering Committee, in which the two public contributors agreed with this decision. 4. To emphasise opportunities for patients to ask questions throughout 5. To make visual updates to the DST, including larger and darker font and use of gender-neutral people in images | 1. Suggested to include a statement/disclaimer at the bottom of the DST, signposting to more detailed information to be accessible via the iFraP website 2. Use of ‘approx’ to demonstrate evidence uncertainty 3. Output of the tool to be developed to allow those having a remote consultation to have an individualised PDF summary of the consultation. Website developed to include additional visuals for those having their consultation on the phone. 4. Reminders throughout tool for the clinician to check patient understanding and provide opportunities for the patient to ask questions 5. Updates made to the visuals of the tool to support reading of text and representation of images |
| Community of Practice meeting 5  (n=2 public contributors, 1 Royal Osteoporosis Society patient advocate. Total of 13 attendees) | To discuss potential updates to the iFraP DST prototype, informed by the feasibility testing findings | Public contributors told us:   1. Concern that ‘managing worry’ might be too broad of a title, and that this conversation should center on worries about fractures 2. They want to know ‘who will I be followed up by, and when?’ | 1. Suggested ‘getting support’ with the inclusion of additional signposting and focus on fracture – such as domestic abuse websites and helplines 2. Opportunity for personalized information about follow up to be entered by the clinician, appearing on the patient’s Personalised Bone Health Record |
| ^1^Hawarden A, Jinks C, Mahmood W, Bullock L, Blackburn S, Gwilym S, Paskins Z. Public priorities for osteoporosis and fracture research: results from a focus group study. Archives of osteoporosis. 2020 Dec;15:1-0.  ^2^Paskins Z, Jinks C, Mahmood W, Jayakumar P, Sangan CB, Belcher J, Gwilym S. Public priorities for osteoporosis and fracture research: results from a general population survey. Archives of osteoporosis. 2017 Dec;12:1-8. | | | |
