## Supplementary Data S1 for "The Michael Mason Prize: Development and feasibility testing of a complex intervention to improve adherence to fracture prevention medicine using a person-centred approach"

### Supplementary Data S1: Additional information about intervention feasibility testing methods and findings

**Methods**

FLS clinicians working in Midlands Partnership University NHS Foundation Trust’s (Staffordshire, England) FLS based in secondary care were invited to take part in the feasibility testing. This FLS site operates a ‘one-stop-shop’ model of care, meaning that, if appropriate, patients have a bone density scan (Dual-energy X-ray absorptiometry [DXA]), nurse assessment, drug treatment recommendation, and blood tests as part of one consultation. FLS clinicians were purposively sampled to gain maximum variation in years of FLS experience and professional role (nurse and allied health professional). If they consented, FLS clinicians were asked to complete the Enhanced Consultation Skills Training Course, including an eLearning package and role play session with other participating colleagues (as described in Table 2). After each component of the training course, clinicians were asked to complete a feedback form.

Patients requiring an FLS appointment invitation, because they had recently had a fragility fracture, were identified by the FLS, as per usual care. Before sending out the appointment invitation, the FLS sent out a study pack, including an invitation letter, participant information sheet, and expression of interest form. Potential participants who provided an expression of interest were contacted by the research team to discuss eligibility and arrange an appointment. If the FLS appointment was face to face, consent was obtained prior to their appointment. If by telephone, consent was obtained remotely, in advance of the appointment.

The feasibility testing was conducted over three iterative testing cycles. Within each cycle, FLS clinicians were asked to deliver the prototype iFraP intervention to a selection of consenting patients. Each consultation was observed and audio recorded by an experienced qualitative researcher (LB, female, applied health services researcher). A predefined fidelity checklist, which detailed use of the DST and training components, was completed.

Immediately after each consultation, the patient completed a semi-structured interview, guided by a topic guide and observations noted in the fidelity checklist. After each testing cycle, the FLS clinician(s) who delivered an iFraP consultation was asked to complete a semi-structured interview. If an FLS clinician delivered consultations in more than one cycle, they had the opportunity to complete multiple interviews. Each FLS clinician interview was guided by a topic guide, with opportunities to explore areas interest from the observed consultation. The prototype iFraP intervention and audio clips of the FLS consultation, were available if required, to facilitate discussions in-interview.

**Findings**

Five FLS clinicians (1 male; 4 female) engaged with the Enhanced Consultation Skills Training Course, with four completing all elements of the course. All four FLS clinicians went onto deliver the prototype intervention in 10 appointments with consenting FLS patients.

Twenty-seven patients were sent a study pack and 13 returned an expression of interest form. When contacted to discuss the study, ten patients agreed to participate and iFraP FLS appointment booked in one of 3 iterative testing cycles (n=3 cycle 1; n=3 cycle 2; n=4 cycle 3). Eight of the consultations were completed in-person, and two completed by telephone.

Each FLS clinician(s) who delivered the prototype iFraP intervention in each cycle were interviewed. In total, the four FLS clinicians completed seven interviews. Demographic characteristics of the four FLS clinicians are not provided to preserve anonymity.

Ten patients received the prototype iFraP intervention and participated in an interview, demographic characteristics are provided below.

| **Table for Supplementary information S1.** Feasibility testing patient demographics | |
| --- | --- |
| **Patient demographics** | **N (%)** |
| Sex |  |
| Female | 9 (90) |
| Age (years) |  |
| 50-60 | 5 (50) |
| 61-70 | 2 (20) |
| ≥71 | 3 (30) |
| Fracture site |  |
| Wrist | 3 (30) |
| Spine | 4 (40) |
| Femur | 1 (10) |
| Upper arm | 1 (10) |
| Drug recommendation |  |
| Recommended drug during FLS appointment | 4 (40) |
| Referred for alternative drug treatment | 3 (30) |
| No drug recommended | 3 (30) |
| Consultation type |  |
| Face to face | 8 (80) |
| Telephone | 2 (20) |

***FLS clinician training feedback***

FLS clinicians (n=5) rated each element of the eLearning as “good” or “very good”. FLS clinicians described the eLearning as ‘simple to use’, ‘easy to understand and follow’. When asked for recommended improvements to the eLearning, two suggested that no improvements were required. The other two clinicians suggested that they wished to see ‘more videos’ of the DST in use and a wish to save eLearning progress.

Three of the four FLS clinicians who completed the role play session submitted a feedback form, also rating each element as “good” or “very good”. FLS clinicians recommended that the structure of the role play session was amended to allow more time for group feedback.

When asked “what changes are you planning to make to your practice?”, free-text responses suggested that they intended to elicit patient beliefs, change their use of osteoporosis terminology, and use evidenced health literacy and risk communication techniques:

*“I might ask and phrase my questions slightly differently going forward and I will choose some different terminologies when explaining patients risks. I will also find out what their knowledge of osteoporosis is at the beginning of my consultation and regularly check to see if they have understood what we have just discussed.”*

*“Lots! regularly checking patients understanding, find out what they already know and what they really want to know and what is important to them.”*

***Intervention fidelity***

All ten appointments were observed, audio recorded, and fidelity checklist completed. Consultations lasted a median of 22 minutes (range 8 – 27 minutes).

In summary, the DST was used in all observed FLS consultations. In the majority of consultations, clinicians asked the patient for their thoughts about their bone health (n=7, 70%), explained the patient’s personal risk factors (n=9, 90%) and how osteoporosis can be controlled using positive language (n=7, 70%), as prompted by the DST. Three patients did not receive a drug recommendation and therefore received a lower dose of the DST (skipping sections related to medicines).

The fidelity checklist also captured whether clinicians implemented elements of the Enhanced Consultation Skills Training Course. The FLS clinician described fracture risk, using simple frequencies in six consultations (60%), used TeachBack in four consultations (40%), and asked the patient if they had any questions in nine consultations (90%).

Five patients (50%) did not receive a copy of the Bone Health Record, because the patient was not recommended a drug treatment (n=3) or printer issues (n=2).

***Interviews***

Interview findings are presented in Table 5.
