## Supplementary Figure S1 for "The Michael Mason Prize: Development and feasibility testing of a complex intervention to improve adherence to fracture prevention medicine using a person-centred approach"

### Supplementary Figure S1: iFraP programme theory

| **Problem:**  Poor fracture prevention treatment uptake |  | **Intervention Resources** |  | **Processes** |  | **Mechanisms** | | |  | **Long-term outcomes** |
| --- | --- | --- | --- | --- | --- | --- | --- | --- | --- | --- |
|  |  |  |  |  |  | **FLS clinician**   - Increased confidence in using DST, communicating risk, health literacy approaches, eliciting and addressing patient preferences and values and SDM - Identifies appropriate treatment options   **PATIENT**   - Greater involvement in decision making - Satisfaction with information/knowledge about condition - Realistic expectations of own fracture risk - Illness perceptions: increased perceptions of coherence and controllability - Medicine perceptions: Fewer doubts about necessity; lower concerns - Increased confidence in decision - Improved decision quality - Increased rates of treatment initiation   **Post-consultation**   - Consistency of messages across primary care and FLS | | |  |  |
|  |  | - FLS - Evidence summaries - DST to depict individualized fracture risk and medicine benefits, harms and practical considerations - Training package - Individualised patient information: the ‘personal BHR’ - Patient information website |  | - FLS clinician receives training in using DST communicating risk, health literacy approaches, eliciting and addressing patient preferences and values and SDM - Trained HCP conducts FLS consultation using DST - Patient given/sent the BHR info. leaflet and signposted to website - GP receives same information |  |  |  |  |  | **PATIENT**   - Persistence with osteoporosis medicine at 3 months - Lifestyle outcomes (physical activity, smoking, alcohol) - *Increased incidence of drug adverse events* - Fracture reduction - Increased onwards appointments, investigations, and referrals   **NHS**   - Guideline adherence - Reduced costs of fracture care |
| **Targeted determinants:**  patient health beliefs |  |  |  |  |  |  |  |  |  |  |
| **Aim:** to improve informed decision making in FLS consultations |  |  |  |  |  |  |  |  |  |  |
| **Context:** Consultations conducted in pre-existing specialist fracture prevention services (Fracture Liaison Service - FLS)  **Contextual factors:** variation in FLS models of care, including whether the consultation is face-to-face or remote, whether the patient has a bone density scan, whether an initial prescription is offered, adherence as a key performance indicator of services, disconnect in advice given to patient between FLS and primary care; media and wider social influences on health behaviours | | | | | | | | | | |
| *******BHR bone health record, DST decision support tool, FLS Fracture Liaison Service, GP general practitioner, SDM shared decision-making  *Italics denote ‘dark logic’ intervention mechanisms/outcomes that may have negative consequences*  **Purple** = key contextual factors identified during intervention development and feasibility testing leading to programme theory updates | | | | | | | | | | |
