## Supplementary Figure S2 for "The Michael Mason Prize: Development and feasibility testing of a complex intervention to improve adherence to fracture prevention medicine using a person-centred approach"

### Supplementary Figure S2: Storyboard to demonstrate iterative development of the iFraP DST

This supplementary material shows how development of the iFraP DST progressed over time, in collaboration with the Community of Practice and public contributors insights and informed by development and testing findings.

1. Iterative development of the ‘your bone health and risk of breaking a bone’ DST page
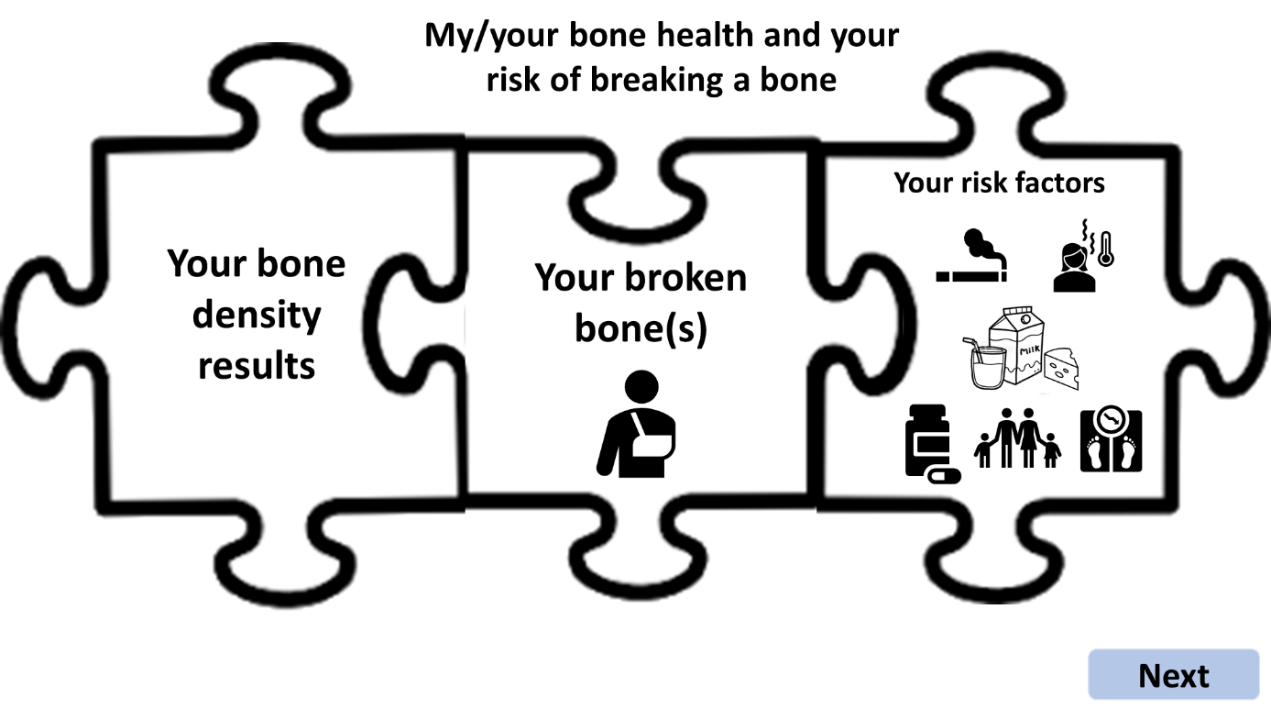

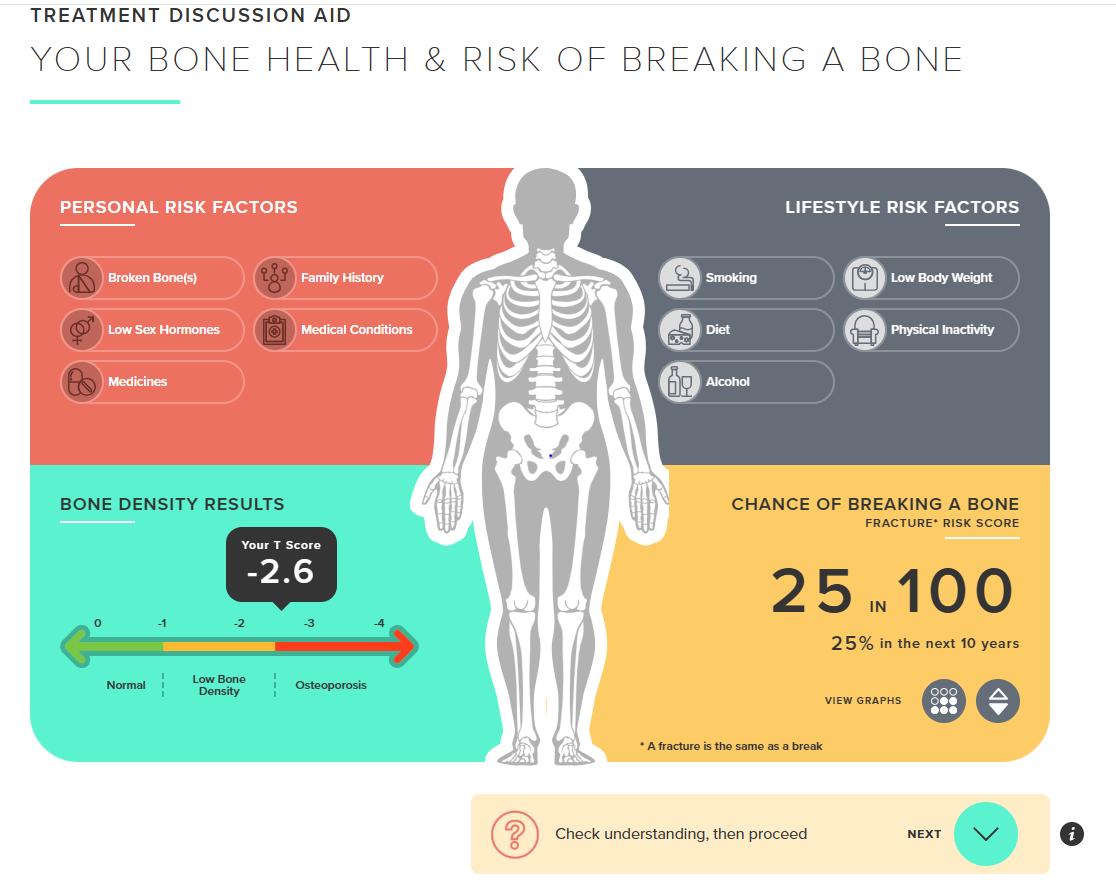

2. Iterative development of the ‘improving your bone health’ DST page

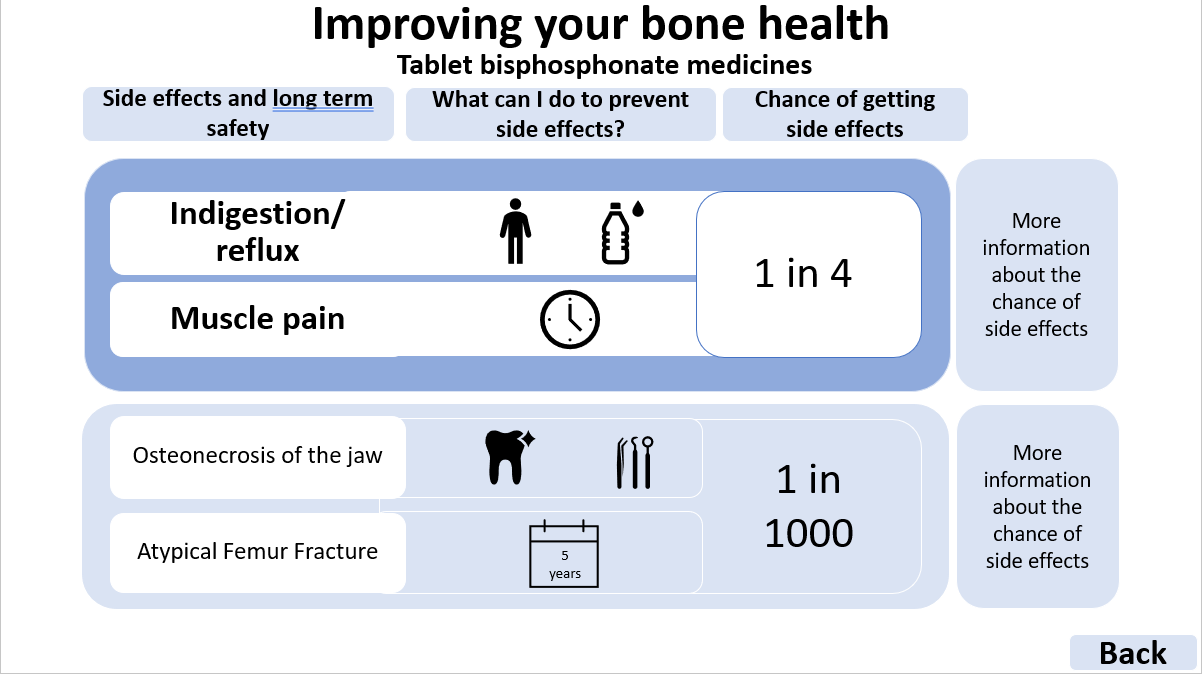

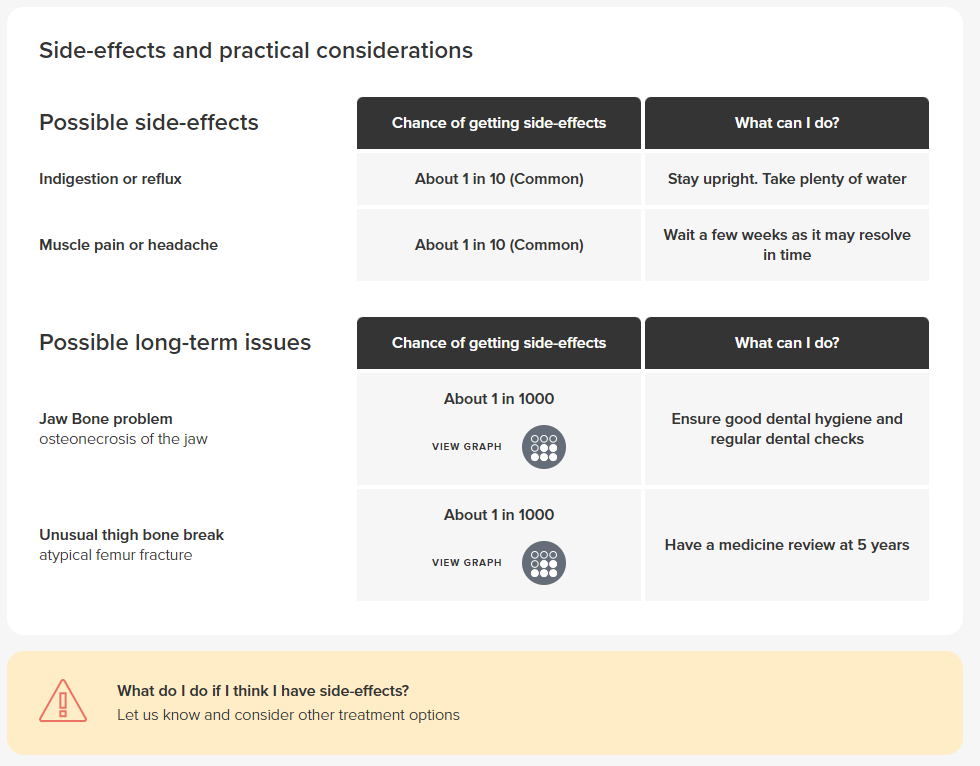

**Developed in collaboration with Prescribing Decision Support Ltd.*
